# Basket Trials in Gynaecology: A Simulation Study Comparing Novel Designs with Current Practice

**DOI:** 10.64898/2026.09.10.26362429

**Authors:** Katie Stocking, James M. S. Wason, Jamie J Kirkham, Andy Vail, Jack Wilkinson

## Abstract

**Background:** Basket trials provide a framework for evaluating an intervention across multiple subtrials within a single protocol. By enabling information sharing, they may improve efficiency and reduce reliance on small, underpowered studies. In gynaecological conditions, we often see patients with the same condition presenting with different symptoms. Although the symptoms are different, treatment is often the same, leading to small and often statistically underpowered trials. Master protocols such as basket trials have the potential to reduce waste in research by allowing the evaluation of a single intervention in multiple disease subtypes. In a hypothetical gynaecological basket trial, this would allow the inclusion of patients with the same underlying condition, but with differing “most bothersome” symptoms, to be included in the same randomised basket trial. In this article, we compare the use of basket trials in gynaecology to typical approaches to trial design and analysis.

**Methods:** Our simulation study evaluates the performance and utility of basket trial designs in hypothetical gynaecological settings. We compare information-sharing strategies treatment effect borrowing (TEB) and treatment response borrowing (TRB) to strategies closely emulating typical gynaecological trial designs: no borrowing (NB, analogous to separate randomised controlled trials) and complete pooling (CP, all patients analysed within a single trial). Performance was measured by key operating characteristics, including coverage of credible intervals, power, credible interval (CI) width, bias, mean squared error (MSE), and empirical standard error. We explore the impact of sample size, treatment effect heterogeneity and control group response, evaluating the balance between efficiency and the risk of misleading inference.

**Results:** Coverage was approximately 95% across all scenarios and methods. CP produced suboptimal coverage under treatment effect heterogeneity and yielded the narrowest CIs in homogeneous scenarios, while NB produced the widest. NB minimised bias under high heterogeneity but incurred the largest MSE, whereas CP exhibited the greatest bias when treatment effects differed across subtrials. TEB generally outperformed TRB in bias and MSE when control arm heterogeneity was present, whereas CP resulted in the highest bias.

**Conclusion:** Adaptive borrowing offers a more robust compromise compared to No Borrowing (independent trials) and Complete Pooling (a single large trial). We conclude cautiously that information borrowing offers efficiency gains, but basket trials require clear clinical and biological justification.

## Background

Patient-centred innovative trial designs are becoming increasingly common in clinical research. These trials, often referred to as master protocols, have the potential to reduce waste in research, by design. Instead of multiple small and often statistically underpowered trials, master protocols expedite intervention development in one large clinical trial. These trial designs either (i) evaluate several interventions in a single disease type (umbrella trials) or (ii) evaluate a single intervention in different disease types (basket trials). Originally developed for use in oncology, in more recent years master protocols have been used in research in other clinical areas such as auto-inflammatory conditions (1), autoimmune and neurological conditions, and HIV (2, 3). Whilst most early basket trials were non-randomised and single-arm, more recent studies explored their utility in later phase, randomised settings (4).

Randomised controlled trials (RCTs) are widely considered the gold standard for the evaluation of interventions. However, in many areas of clinical research, including gynaecology, conventional RCTs can be inefficient. Gynaecological conditions such as Polycystic Ovary Syndrome (PCOS) and endometriosis are characterised by substantial heterogeneity in symptom presentation, with individuals often experiencing different combinations of symptoms and placing different importance on those symptoms. Despite this, eligibility criteria and primary outcomes are frequently defined according to a single symptom, excluding patients whose most bothersome symptom differs despite having the same underlying condition. Our previous systematic review demonstrated that this is common in gynaecological research, with over half of PCOS trials (55%) and almost half of endometriosis trials (46%) restricting eligibility based on symptom presentation, while many trials measured outcomes that were not relevant to all participants (38% and 31%, respectively) (5). This exacerbates research waste by limiting recruitment, reducing the applicability of trial findings, and making evidence synthesis challenging because different studies evaluate different patient populations and outcomes.

Basket trial designs offer a potential solution to these challenges by allowing participants with different predominant symptoms to be enrolled within a single protocol and allocated to symptom-specific subtrials while receiving the same intervention. This approach is particularly appropriate for gynaecological interventions that target the underlying disease rather than an individual symptom, increasing research inclusivity by allowing a broader range of patients to participate while generating evidence across multiple patient-important outcomes. Furthermore, the common underlying disease and shared intervention suggest that treatment effects across symptom-specific subtrials are unlikely to be entirely independent. Borrowing information across these related subtrials may therefore increase statistical efficiency and improve the precision of treatment effect estimation, particularly where individual symptom groups contain relatively few participants. Evaluating the performance of such borrowing approaches is therefore an important step towards developing efficient basket trial methodology for gynaecological research, and facilitating more inclusive and patient-centred trial designs.

The analysis of basket trials centres around the heterogeneity of patient outcomes within the trial. In general, either: patient outcomes are considered heterogeneous and analysis of subtrials is carried out separately (no borrowing); patient outcomes are considered homogeneous and are analysed as one pooled data set (complete pooling); or patient’s potential outcome heterogeneity is accounted for measured using an appropriate analysis strategy. Borrowing information uses data from related patient groups to improve the precision of treatment effect estimates. Adaptive borrowing methods allow the extent of information borrowing to depend on the similarity between groups, typically borrowing more when treatment effects are alike and less when they differ. There have been several analysis methods developed for accounting for potential heterogeneity in basket trials (6–12). These statistical methodologies differ primarily in how they determine the amount of information to share between subtrials. This paper builds on work by Zheng and Wason (3), and Ouma et al (13), which proposed a commensurate predictive prior (CPP) approach to model the commensurability of treatment effects, and groupwise mean responses to permit the borrowing of information across complementary subtrials. The aim of this study is to compare the use of basket trials employing these modelling strategies, to a typical approach to trial design in which participants with different symptoms are recruited to separate, small trials and analysed separately.

## Methods

### Aim

Our simulation study aimed to evaluate the use of basket trials in a gynaecological setting by comparing no borrowing (e.g. individual randomised controlled trials), complete pooling (e.g. pool-all patients in one trial), treatment effect borrowing (TEB) and treatment response borrowing (TRB).

### Data Generating Mechanisms

We simulated a randomised basket trial setting in which patients are recruited and placed into subtrials depending on their “most bothersome symptom” (MBS) in a typical gynaecological setting. Figure 1 illustrates a hypothetical randomised basket trial in gynaecology, where there are three identified MBS.

**Figure 1.**
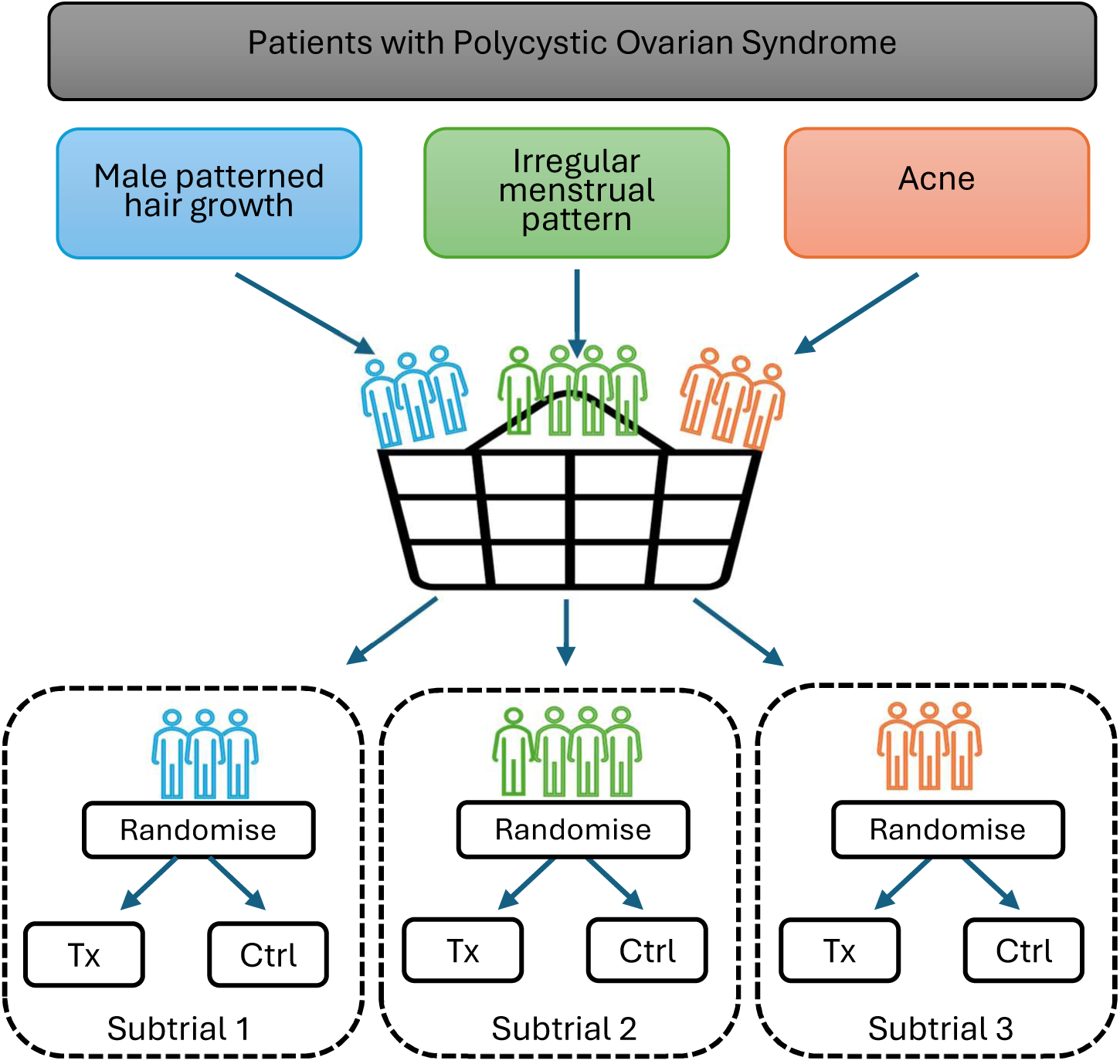
**Illustrative depiction of a randomised basket trial in gynaecology**

Our simulation was informed by the findings from our systematic review of current practice in systematic reviews and their component trials in gynaecology (5). Our chosen primary outcome of the simulated basket trial was a continuous visual analogue scale which will be used to evaluate the impact of the participant’s most bothersome symptom, measured at baseline and end of trial and calculated as a change score (with more positive change scores being better). This outcome was the same in each subtrial.

In order to calculate the required number of simulation replications needed to control the Monte Carlo standard error (MCSE) of the estimated power, we used guidance by Morris et al (14). We calculated that 3600 replications would estimate a true power of 50% with an MCSE of 0.0083. We selected 50% to be conservative – if power is higher or lower, the MCSE will be smaller. With a power of 50% and an MCSE of 0.0083, this gives an approximate expected 95% CI width of ±1.6 percentage points.

### Method of simulating outcome data

For all scenarios, we simulated data from hypothetical randomised basket trials with *j* subtrials, and *i*=1,…,*n*_j_ patients in each subtrial *j* = 1, …, *J*. Patients were randomised into two equal groups (treatment or control) within each subtrial. The binary treatment variable *T*_ij_ split the observations in each subtrial into two equal groups, 0 (control) and 1 (treatment) such that *T*_ij_ = 1 if patient *i* in subtrial *j* receives the treatment and *T*_ij_ = 0 if the patient receives the control.

In order to simulate the observed continuous clinical outcome (*y*_ij_) in each subtrial we modelled:

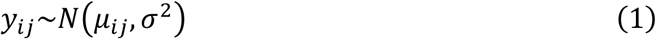

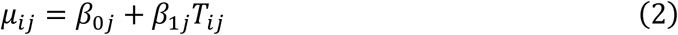

Where *β*_1j_ is the subtrial specific treatment effect and *β*_0j_ is the groupwise mean response, with an inter-patient SD fixed at *σ* = 1. In the Bayesian analyses, we specified a random effects model for 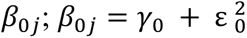. A weakly informative normal prior was specified for γ_0_, γ_0_ ∼ N(0,5) and 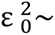∼ Half Normal(1), where Half Normal(1) denotes a truncated N(0,1) to cover the range (0,∞) (13).

We considered hypothetical randomised basket trials with *j* = 7 subtrials. We chose 7 subtrials as this represents the number of categories of outcomes found in our systematic review in PCOS (Fertility and Ovulation, Menstruation, Body composition, Hirsutism, Acne, Mental health and Quality-of-life) (5).

### Scenarios

We considered scenarios varying the overall sample size of the basket trials, in addition to the treatment effect sizes and the mean response in the control arm for the individual subtrials.

Three overall sample size configurations were selected based on previous basket trial designs and findings from our systematic review to reflect small, medium, and large trial scenarios. In our systematic review, we identified 89 PCOS trials using various strategies to measure and analyse the participant’s outcome(s). For each outcome category, we extracted the minimum, 25^th^ percentile, median, 75^th^ percentile and maximum number of randomised participants. To ensure the simulated basket trials reflected realistic sample sizes observed in the literature, the sample size assigned to each symptom-specific subtrial was based on the minimum, median, and 75th percentile of trials evaluating the corresponding outcome. Consequently, we assigned the seven subtrials unequal numbers of participants to be randomised (*n*_j_) in to reflect the variation in the trial sizes observed across symptom-specific studies (Table 1).

**Table 1.**
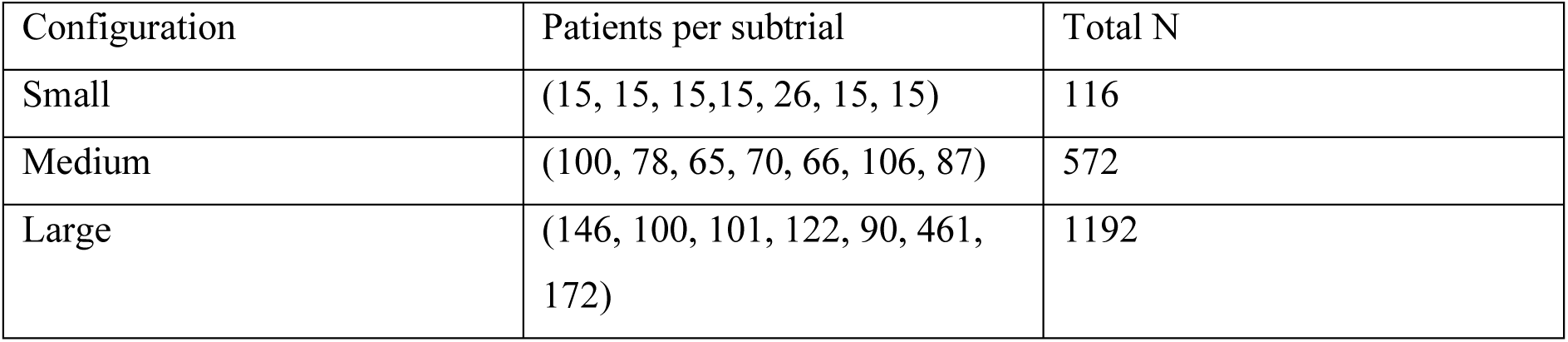
Sample size configurations for simulation.

For each sample size scenario, we examined the impact of varying treatment effect on the performance measures of the basket trial. We varied the treatment effects and mean responses in the control arms of the subtrials, creating 8 different scenarios. Scenario 1 represents the “null” scenario, with no treatment effect in any of the subtrials. Scenario 2 has consistent treatment effects in each of the subtrials. Scenario 3 and 4 represent basket trials high heterogeneity in the subtrial treatment effects (scenario 3), and low heterogeneity (scenario 4). In scenario 1-4 the control group mean was fixed across subtrials. In scenarios 5-8 control group means varied across the subtrials (Table 2). In total, with the three sample size configurations, we will have 24 different sample size-scenario configurations.

**Table 2.**
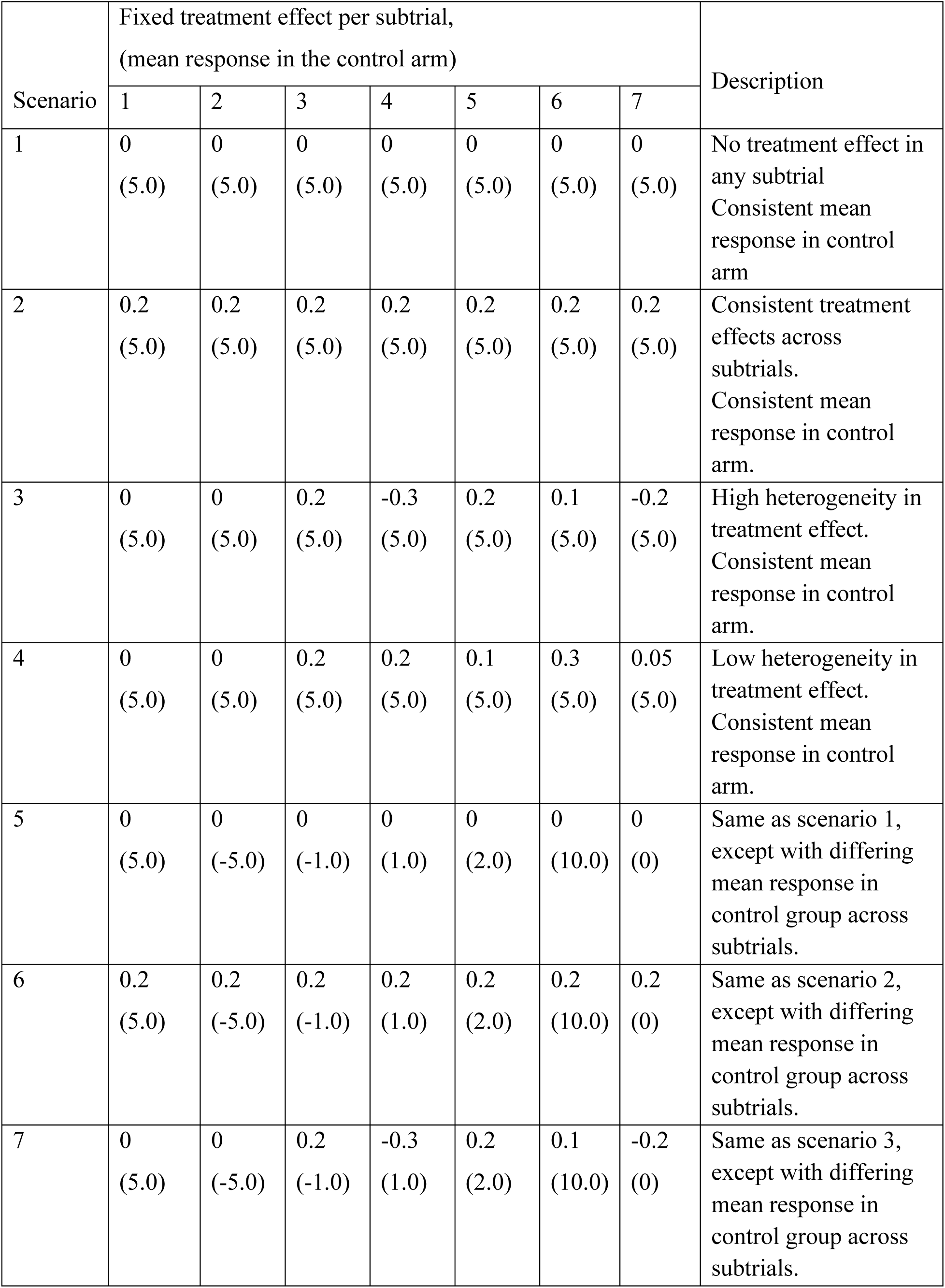
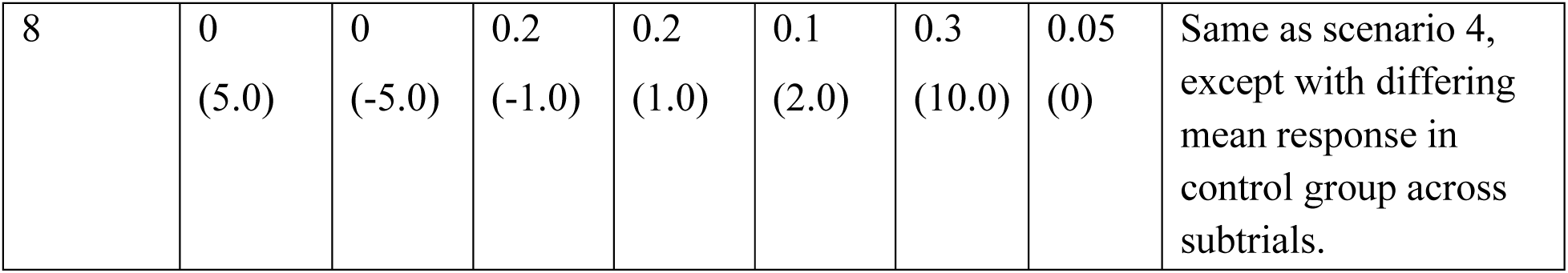
Descriptions of simulation scenarios in relation to the treatment effect and control group mean response.

**Table 3.**
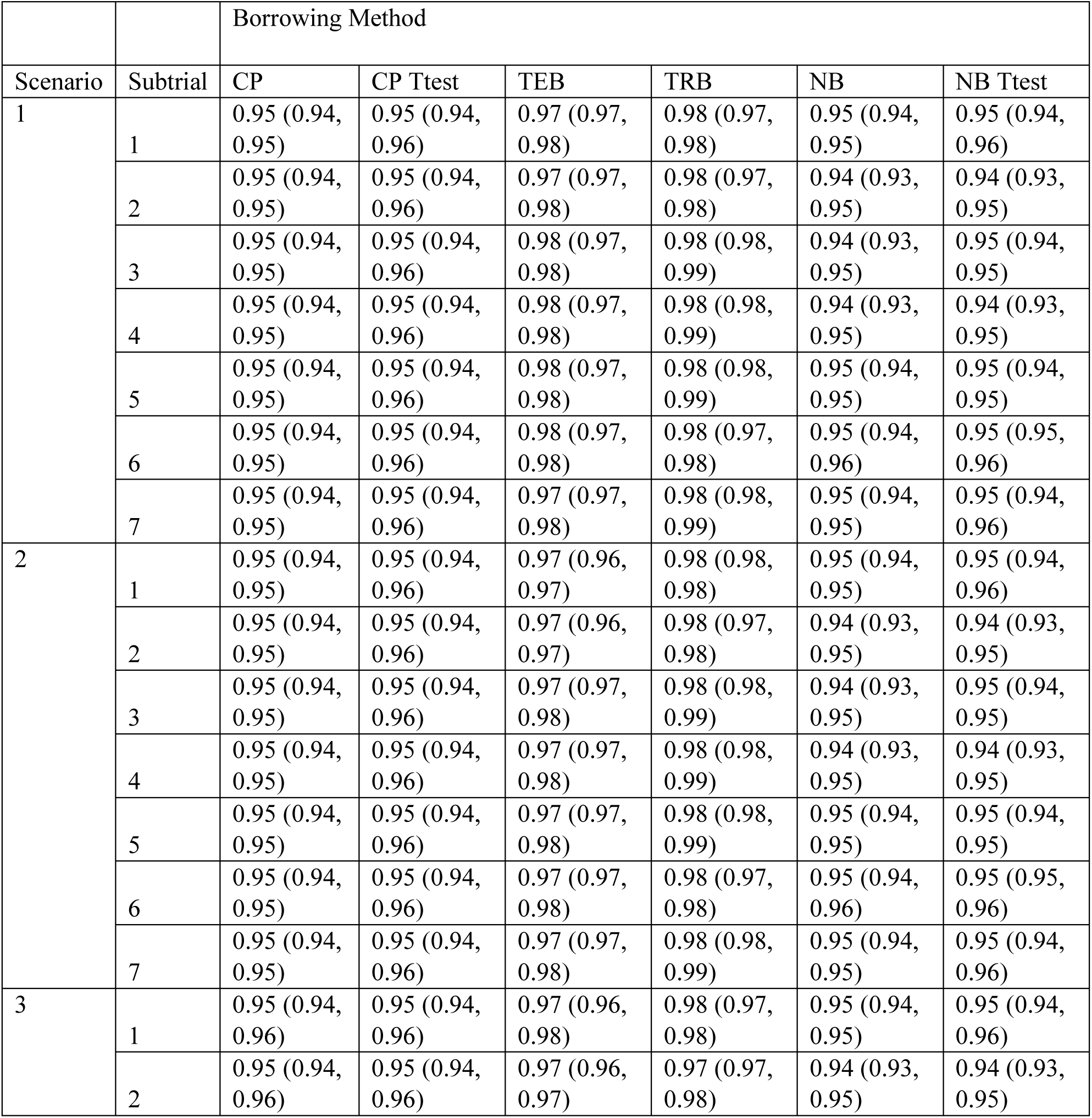
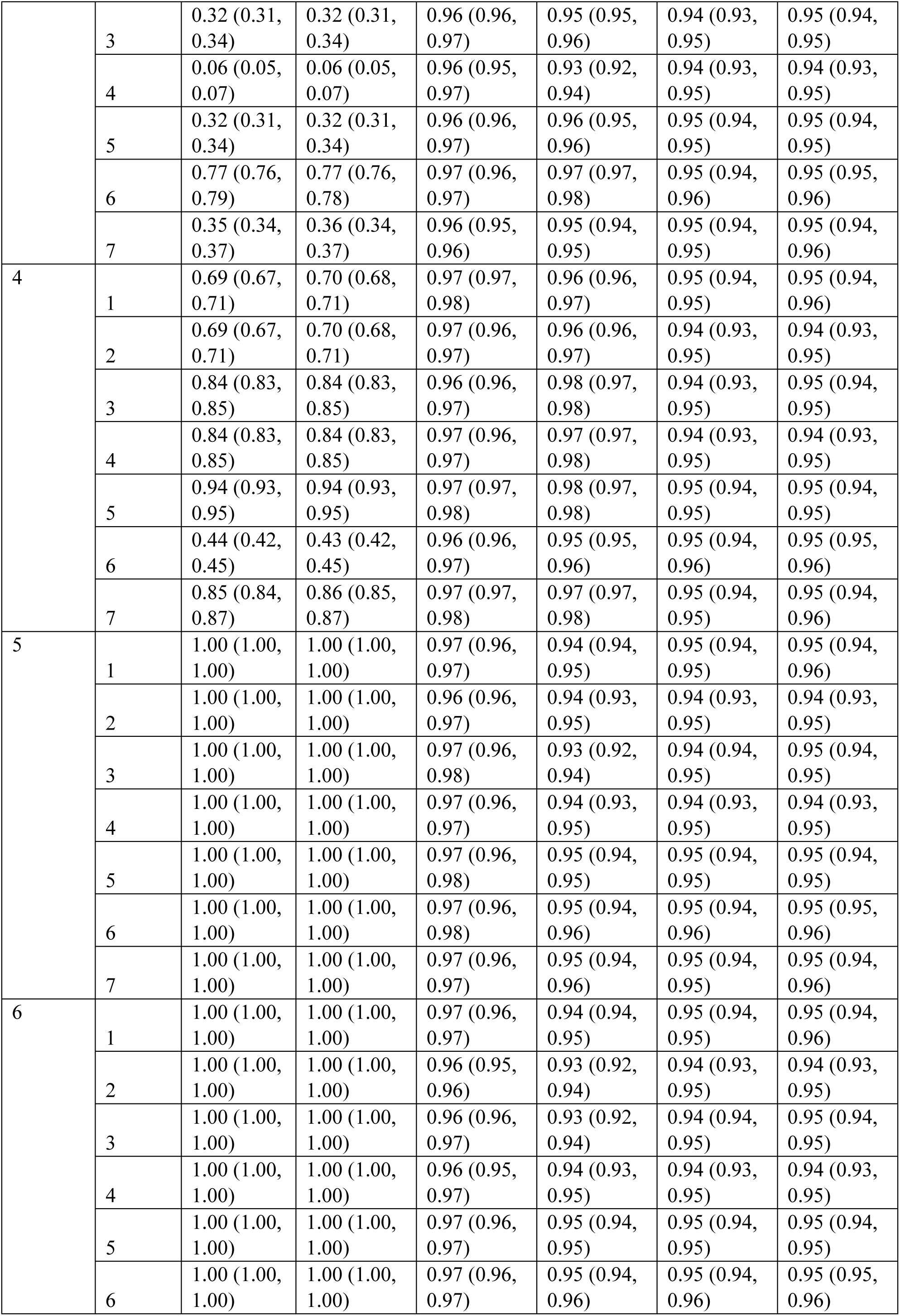
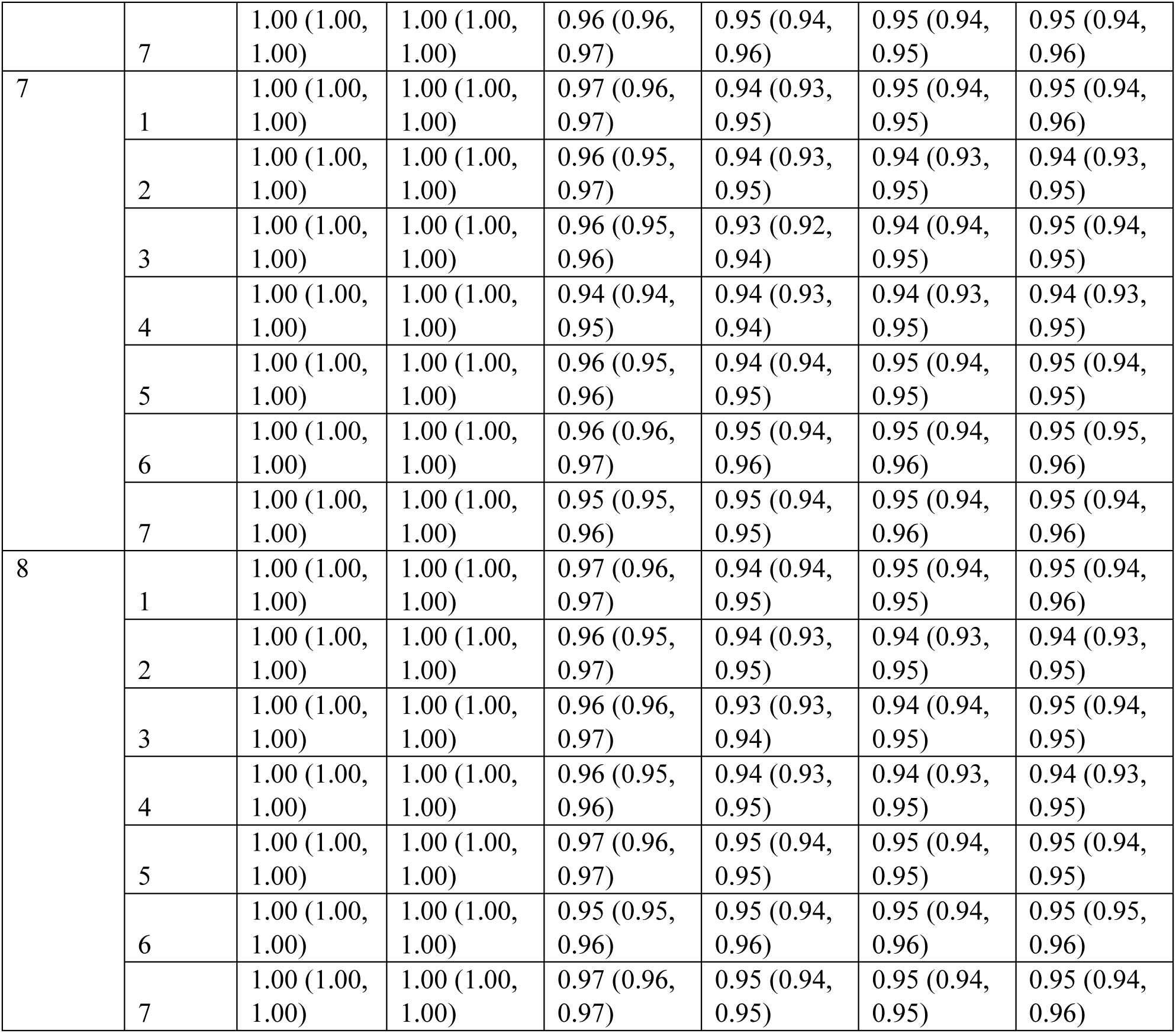
A Coverage with 95% CI for each borrowing method

## Estimand

We estimated the treatment effects as the difference in mean outcome in each subtrial, along with the 95% credible interval.

### Treatment Effect Borrowing and Treatment Response Borrowing

We used the commensurate predictive prior (CPP) approach to model the commensurability of the treatment effects and responses per treatment arm between subtrials in order to leverage information, building on work by Zheng and Wason (3). We evaluated the commensurability between the subtrial-specific treatment effect *β*_1j_ and *β*_1j∗_ where *j* and *j*^∗^ are individual subtrials and *v*_jj∗_ is the precision parameter assessing the degree of consistency between subtrials *j* and *j*^∗^ :

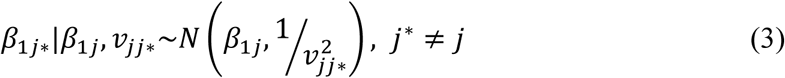

Large values of *v*_jj∗_ indicate strong agreement between subtrial treatment effects and therefore greater borrowing, whereas small values allow the treatment effect in subtrial *j* to differ from subtrial *j*^∗^. We followed guidance by Ouma et al, placing a spike and slab prior on each precision parameter *v*_jj∗_ defined as locally uniform between two limits 0 ≤ α_1_≤ α_u_ (the slab component), and with a probability mass concentrated at a point S >α_u_ (the spike component) (13). The spike and slab prior allowed subtrials with similar treatment effects to receive larger borrowing weights, whereas dissimilar subtrials contributed less information. The implementation of a spike-and-slab prior avoids the assumption of exchangeability between the treatment effects across the subtrials, instead, it uses a distributional discrepancy measure to allow information to be shared only between the more commensurate subtrial(s) (15).

In order to quantify a prior for the commensurability between subtrials, we followed guidance by Zheng and Wason and used the same Hellinger distance (*d*_H_) as quantified by Ouma et al (13, 15):

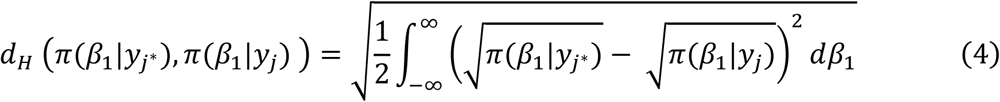

Where *π*(*β*_1_|*y*_j∗_) and *π*(*β*_1_|*y*_j_) are the posterior distributions for subtrials *j* and *j*^∗^. The Hellinger distance provides a measure of the similarity between the posterior distributions of two subtrials and is used to determine the extent of information borrowing. When *d*_H_ (*π*(*β*_1_|*y*_j∗_), *π*(*β*_1_|*y*_j_)) approaches 1, the subtrials exhibit greater incompatibility, leading to increased allocation of probability mass to the slab component of the spike-and-slab prior. This corresponds to a smaller precision parameter (larger variance), thereby reducing the influence of the complementary subtrial. Conversely, when *d*_H_ (*π*(*β*_1_|*y*_j∗_), *π*(*β*_1_|*y*_j_)) approaches 0, this indicates greater similarity between subtrials, resulting in greater weight assigned to the spike component, a larger precision parameter *v*_jj∗_, and increased borrowing of information. The resulting Hellinger distances are subsequently normalised to obtain borrowing weights that determine the contribution of information from each complementary subtrial.

This work was further modified by Ouma et al (13) to extend to modelling the commensurability of the groupwise mean responses (the mean response in either the control or treatment arm within a subtrial) allowing for TRB. Although we implement borrowing by treatment response based on φ_tj∗_, our interest lies on the subtrial-specific treatment effect defined as *β*_1j_ = φ_Ej_ − φ_Cj_ for subtrial *j*, where *t* = *E*, *C* index the treatment and control groups, respectively. We used CPP once more to facilitate information borrowing using a normal predictive prior φ_tj_, with a precision parameter 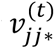. Our normal predictive distribution and Hellinger distance discrepancy measure is taken from Ouma et al and can be described as (13):

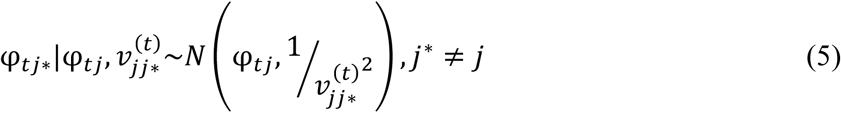

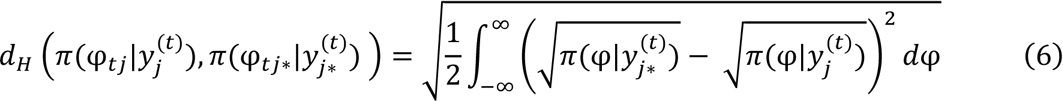

Specifically, our Hellinger distance measure allowed us to utilise information from a complementary subtrial as *d*_H_(φ_tj_, φ_tj∗_) → 0 (high commensurability) and discount information when *d*_H_(φ_tj_, φ_tj∗_) → 1 (high incommensurability).

We used these methods, in comparison to stand-alone analyses (no borrowing and complete pooling in both frequentist and Bayesian frameworks) in order to test the functionality in a theoretical gynaecological basket trial design.

Treatment Effect Borrowing (TEB) considers how similar the treatment effects are across the subtrials, and Treatment Response Borrowing (TRB) considers how similar the responses per treatment arm are (i.e. separately considering each arm). Both approaches allow for the complete discarding of information if the treatment effect/groupwise mean response in subtrials are incommensurate. The CPP approach is of particular use where the sample size within the subtrials is small, making the estimates less precise.

### No Borrowing and Complete Pooling

For both no borrowing and complete pooling, we implemented both a Bayesian and frequentist analysis of the basket trial.

No borrowing treats each subtrial as an independent randomised controlled trial, therefore, each subtrial was analysed in isolation without sharing information across subtrials. The frequentist approach to no borrowing between subtrials used independent t-tests for each subtrial. For the Bayesian approach for patient *i* in subtrial *j*, we used the same data generating models in (1) and (2). An independent Normal prior was assigned to each subtrial-specific treatment effect *β*_1j_ ∼ *N*(0, 10^2^) and intercept *β*_0j_∼ *N*(0, 5^2^).

Complete pooling assumes full statistical exchangeability among all patients in all subtrials. In this scenario, patient data is analysed as it if is from a single randomised controlled trial under the assumption that all trials have identical treatment effects. Both frequentist and Bayesian analyses were conducted. For the frequentist approach to complete pooling, we used a single t-test which pooled all patients across all subtrials. For the Bayesian approach, we modelled:

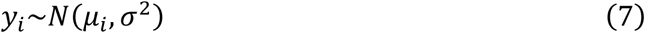

With

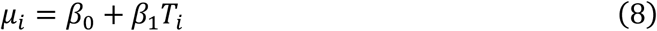

Where *β*_0_ is the overall control group mean, and *β*_1_ is the common treatment effect shared across all subtrials. An independent Normal prior was assigned to the common treatment effect *β*_1_ ∼ *N*(0, 10^2^) and intercept *β*_0_∼ *N*(0, 5^2^). Thus, all subtrials contributed jointly to the estimation of a single common treatment effect.

### Performance Measures

We investigated the utility of basket trial design and analyses using the following performance measures: coverage probability (target: 0.95), frequentist power (true positive rate) and type 1 error (false positive rate), credible/confidence interval width, bias, mean squared error (MSE) and empirical standard error. Coverage probability is the proportion of times the true treatment effect is contained within the 95% credible/confidence interval. Frequentist power was calculated as the proportion of iterations in which the credible/confidence interval excluded the null value (0). In scenarios containing subtrials with null treatment effects, the proportion of credible/confidence intervals excluding zero was interpreted as subtrial-specific Type I error (false positive rate). For subtrials with non-zero treatment effects, this was interpreted as the subtrial-specfic power. Credible/confidence interval width is the average width of the interval estimate, reflecting the precision of the treatment effect estimate. Bias is the mean difference between the estimates and the truevalue. MSE reflects both how far estimates are from the true value and how variable they are. Empirical SE is the observed variability in treatment effect estimates across repeated simulations.

## Results

All code used in this study for both R and JAGS is available at https://github.com/katiestones/basket-sim We describe the performance measures for a medium sample size basket trial, with small and large sample size results reported in the Supplementary Materials. We note that although Monte Carlo 95% CIs were computed for each performance measure, they were extremely precise relative to the point estimates and therefore were not visually discernible in the plots.

### Power (true positive rate) and Type 1 error (false positive rate)

Scenarios 1 and 5 were used to evaluate Type I error since all treatment effects were null, whereas scenarios 2 and 6 evaluated power under homogeneous non-null treatment effects (Figure 2). Scenarios 3, 4, 7, and 8 contained a mixture of zero and non-zero treatment effects in subtrials and therefore allowed simultaneous assessment of subtrial-specific Type I error and power.

**Figure 2.**
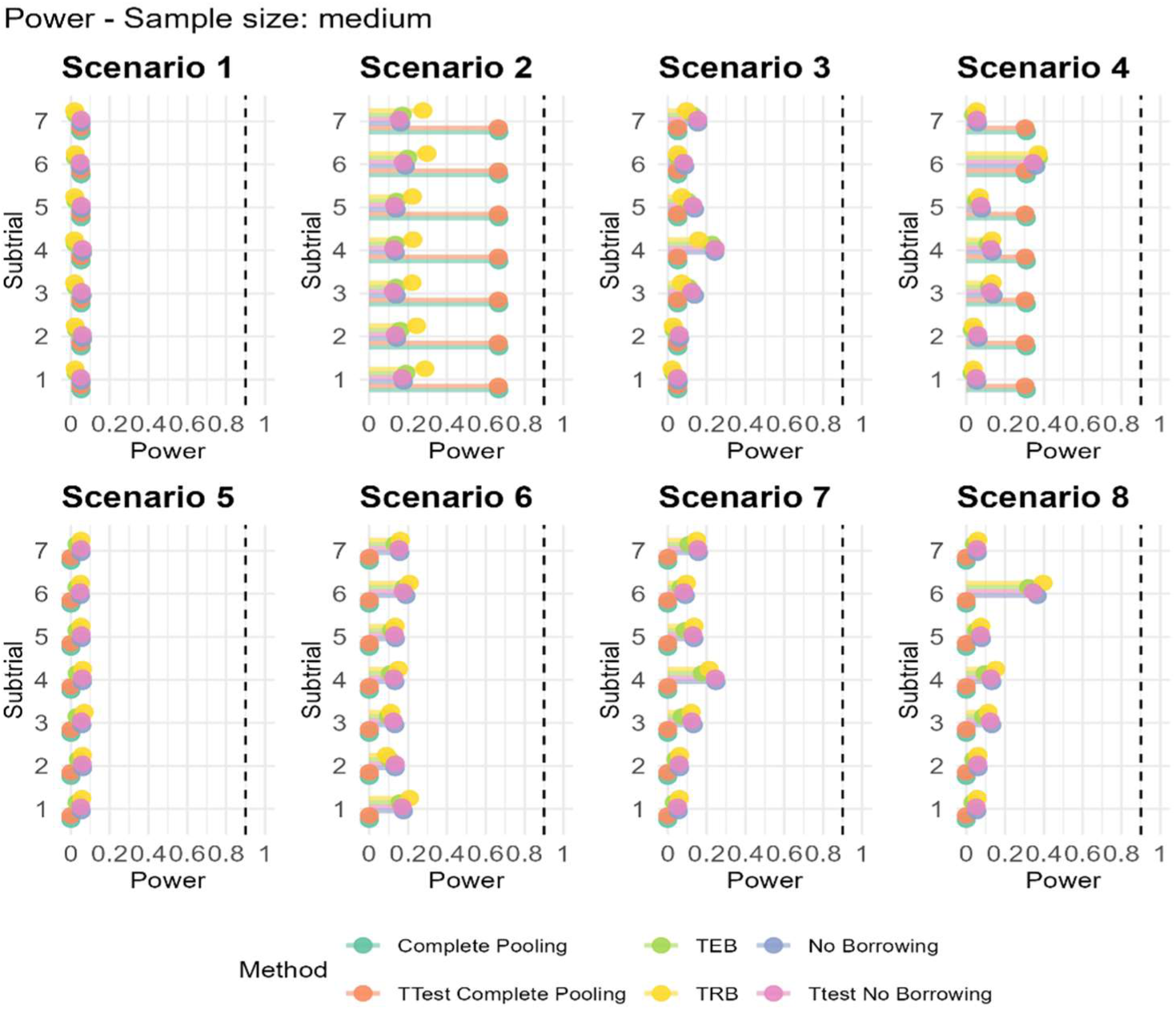
**Type 1 error (scenarios 1 and 5) and Power (all other scenarios) for medium sample size**

In scenarios where the true treatment effect was zero for all subtrials (scenarios 1 and 5), the NB approaches achieved adequate empirical Type I error control across subtrials. The TEB and TRB approaches were more conservative in scenario 1, with Type I error rates of approximately 2-3%, whilst rates in scenario 5 were closer to the nominal 5% level. Complete Pooling maintained Type I error rates close to 5% in scenario 1; however, it was highly conservative in scenario 5, with rejection probabilities equal to zero. In scenarios 2 and 6, where all subtrials had nonzero treatment effects, statistical power remained low. Where treatment effects were homogeneous (scenario 2), CP achieves the highest power. In the case of scenario 6, where treatment effects were homogeneous but the mean response in the control group differed between subtrials, TEB and TRB performed similarly to NB, achieving substantially higher power than CP. In scenario 4 and 8, where we have high heterogeneity in the treatment effects, we saw that in subtrials where the treatment effect is higher (subtrial 6) TEB and TRB, respectively, achieved the highest power.

### Coverage

Across scenarios, coverage was generally maintained at the nominal 95% level or higher for most methods (Figure 3). Scenarios 3 and 4 display the largest differences in coverage between the subtrials when CP is used. This is because CP pools into one estimate, giving an average for *β*_1_. Therefore, the more heterogeneous the true treatment effects are in the subtrials, the further the overall posterior estimate is from the subtrial-specific effects, causing poor coverage. The heterogeneous mean responses in scenarios 5,6,7 and 8 increase variance in the outcome. The improvement in coverage for scenario 7, compared to scenario 3, indicates that control group heterogeneity influenced the coverage of the treatment effect estimates.

**Figure 3.**
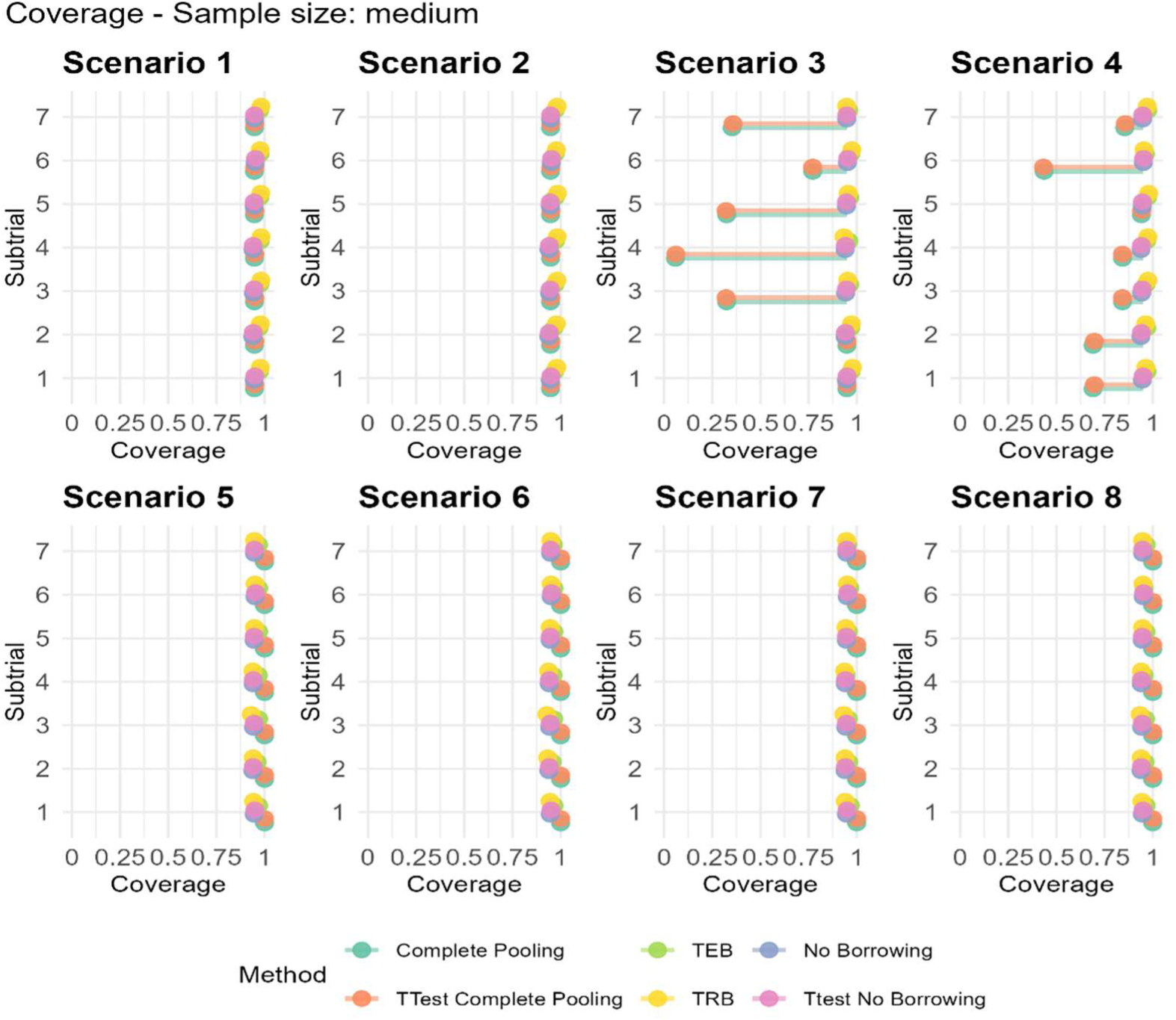
**Coverage for medium sample size**

### Credible Interval Width

Figure 4 shows the mean credible interval (CI) widths for the treatment effect estimates. In Scenarios 1-4, where the mean response in the control arm was consistent across subtrials, CP methods produced the narrowest CIs, followed by TEB and TRB. In contrast, NB methods yielded consistently wider intervals. Introducing heterogeneity in the control arm (Scenarios 5-8) widened the CIs across all methods, in particular, CP methods. In these scenarios, TEB produced the narrowest CIs, followed by TRB and NB.

**Figure 4.**
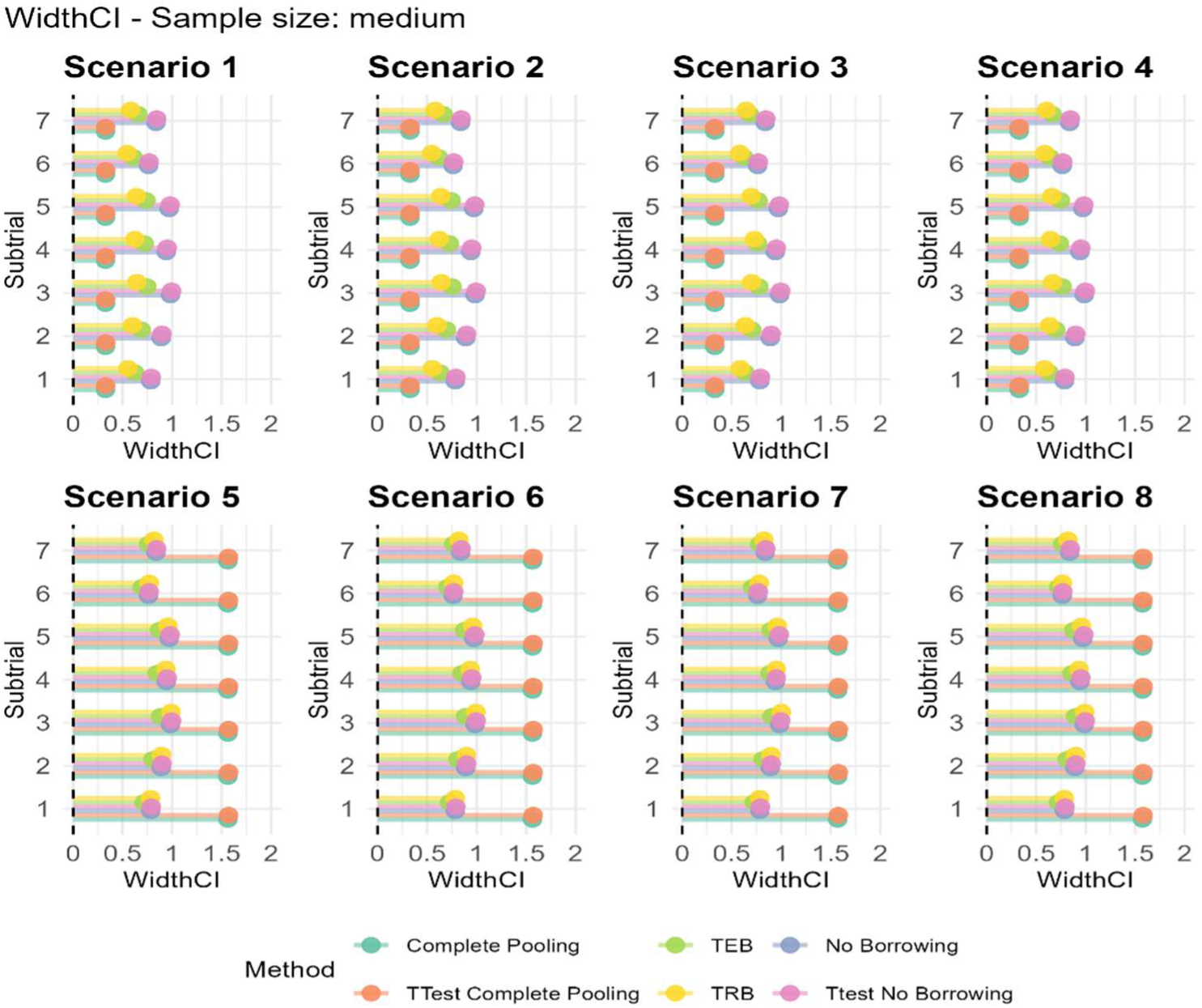
**Width of CI for medium sample size**

### Bias and Mean Squared Error

Figure 5 shows bias and Figure 6 shows Mean Square Error (MSE). In scenarios 1-4, all of the subtrials had consistent mean response in the control group, with differing levels of treatment effects. When the treatment effect and control group response mean is consistent across all methods (scenario 1 and 2), the bias is small across all methods. Notably, the MSE is larger with no borrowing, compared with TEB and TRB. When we have high heterogeneity in the treatment effect (scenario 3) and therefore anticipate limited borrowing of information, no borrowing produces the smallest bias, but the largest MSE. In scenario 3, where the treatment effect is non-zero in subtrials 3-7, complete pooling is associated with a much larger bias than other methods. Moreover, in this case, compared with TRB, TEB produces a smaller bias when there is high heterogeneity in the treatment effect but a slightly increased MSE. When there is low heterogeneity in the treatment effect (scenario 4), the results follow the same trend. In both scenarios, when heterogeneity in the treatment effect is present, no borrowing produces minimal bias but a much larger MSE than other methods. Introducing differing mean responses in the control arm (scenario 5 and 6) we see that TRB has a higher bias and MSE than the other methods. For scenario 7, where treatment effect and response are different both quantitatively and qualitatively, TEB and TRB are comparable in bias, however, TEB produces smaller MSE.

**Figure 5.**
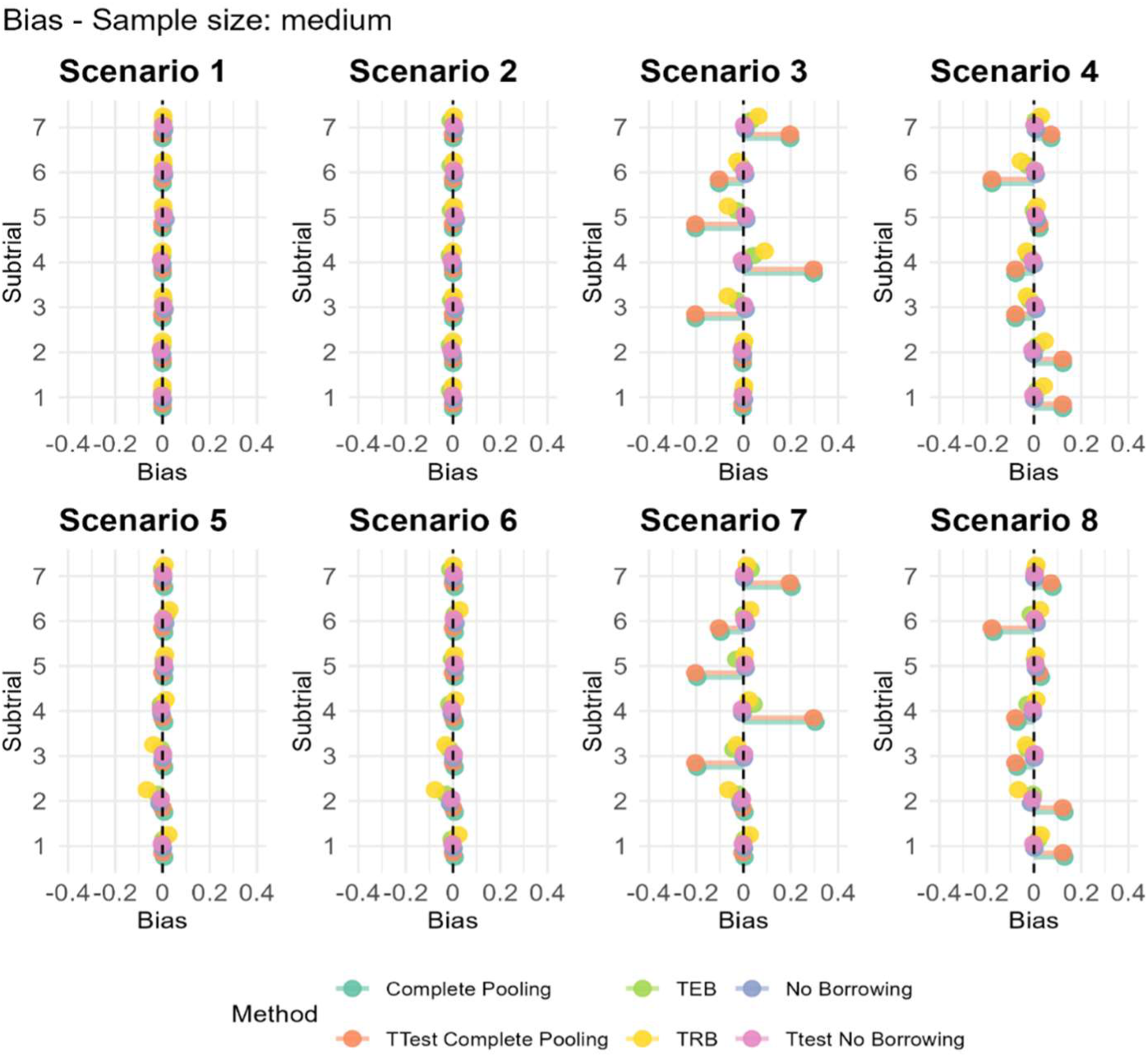
**Bias for medium sample size**

**Figure 6.**
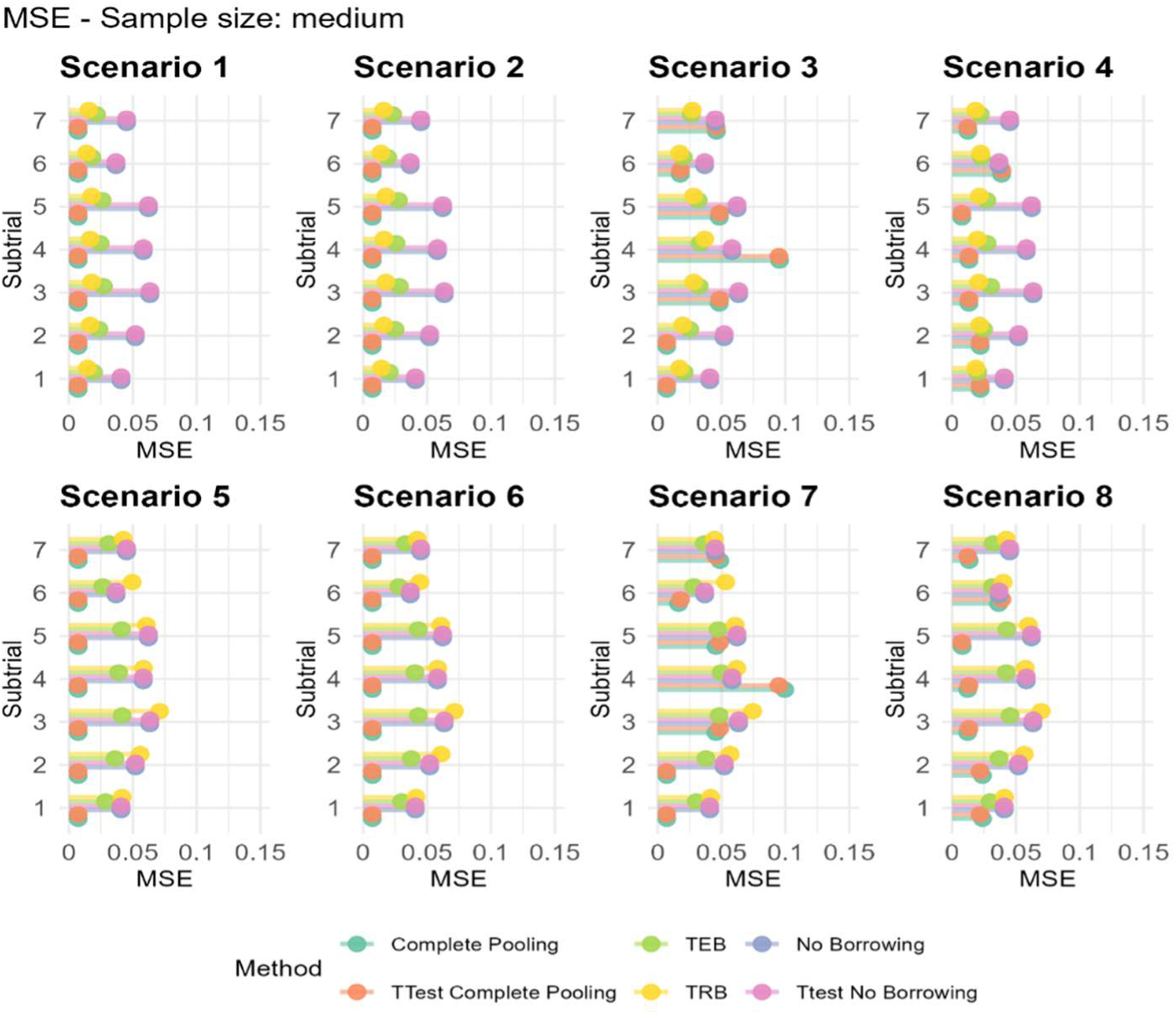
**MSE for medium sample size**

### Empirical Standard Error

Empirical SE was found to be broadly comparable between subtrials within each scenario as the SD and sample sizes were fixed in each subtrial. Differences occurred, however, across methods, with CP having a lower empirical SE than the other methods, as expectedly, pooling reduced variability in the estimate. In Scenarios 1 to 4, where the control group mean remained the same in each subtrial, we see that NB has the highest empirical SE, with TRB performing slightly better than TEB. The opposite is true when the control group mean differs in scenarios 5 to 8, with TRB having a higher empirical SE than TEB. We note that when the control group mean varied in scenarios 4-8, the empirical SE increases for TEB and TRB methods.

### Sample size differences

Using smaller and larger sample sizes to assess performance measures relative to a medium sample size showed that the overall conclusions regarding the comparative performance of the methods remained unchanged.

Larger sample sizes followed the same trends observed for small and medium samples, but with increased power in scenarios 4 and 8, where treatment effect heterogeneity was high, particularly in subtrial 6 with a larger treatment effect (Fig. 8A & Fig. 9A). Coverage conclusions were also unchanged, although for smaller sample sizes the difference between CP and the other methods was less pronounced in scenarios 3 and 4 (Fig. 10A & Fig. 11A). Similarly, larger sample sizes led to much smaller differences in interval widths between methods compared with the small and medium sample settings (Fig 12A & Fig. 13A). As expected, smaller sample sizes increased both bias and mean squared error (MSE), whereas larger sample sizes reduced them, without changing the relative performance of the methods (Fig 14A-17A). Empirical standard errors were largely unaffected by sample size, aside from the expected reduction when larger samples were used (Fig. 18A & Fig. 19A). Overall, although changing the sample size influenced the magnitude of the performance measures, particularly improving performance for larger samples, it did not alter the overarching conclusions, and differences between methods became less prominent as the sample size increased.

**Figure 7.**
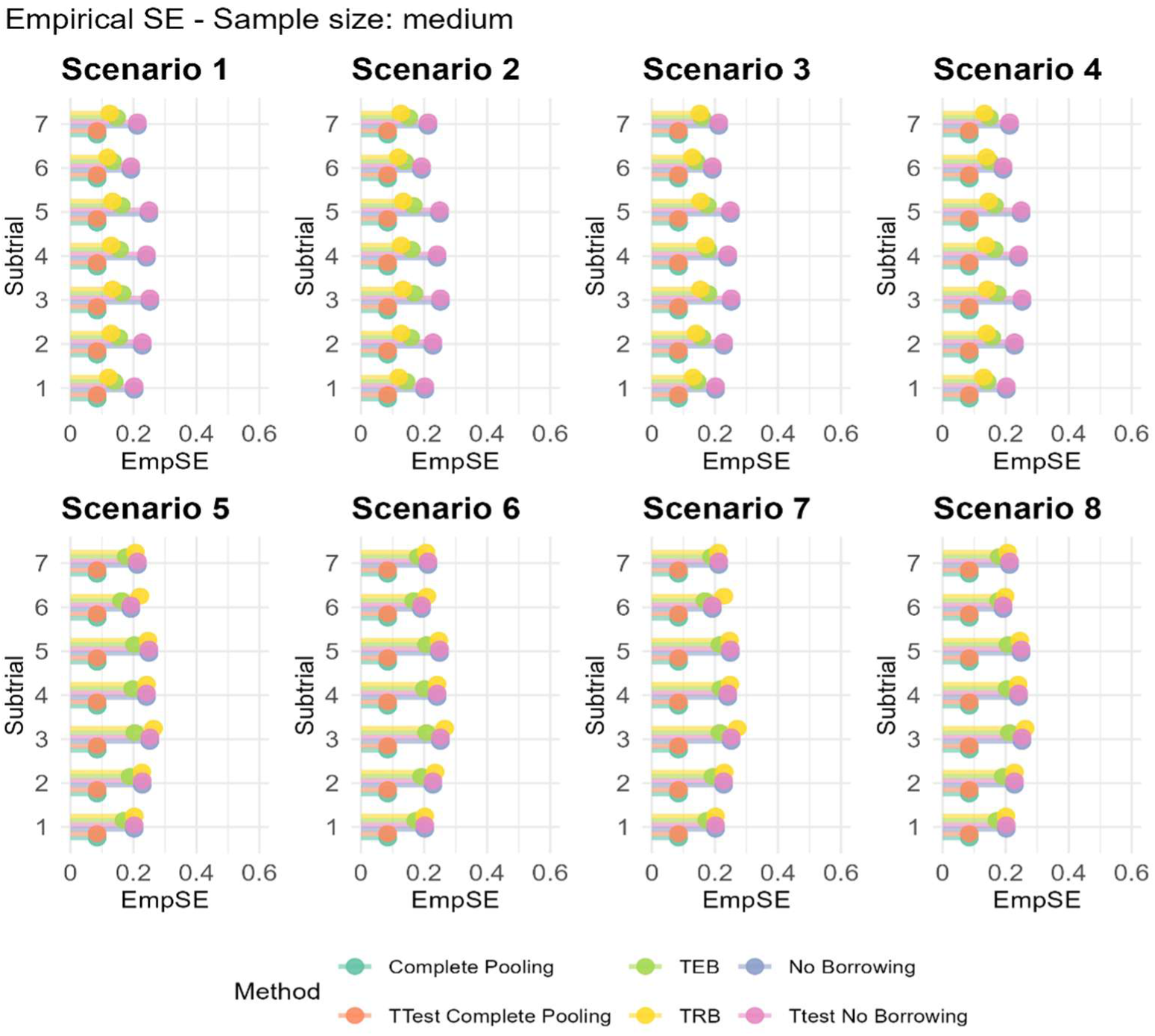
**Empirical SE for medium sample size**

**Figure 8.**
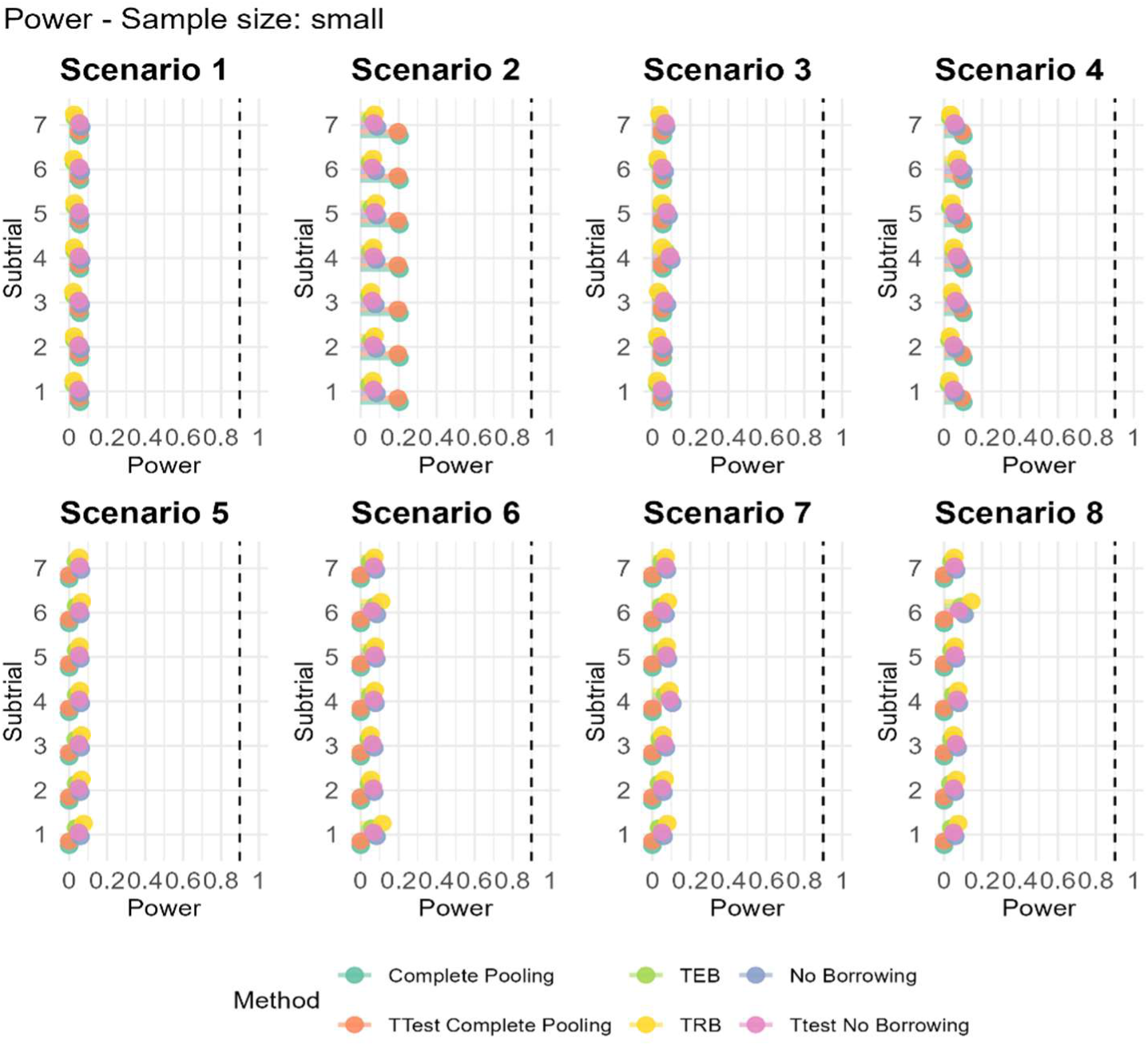
**A Power and Type 1 error for small sample size**

**Figure 9.**
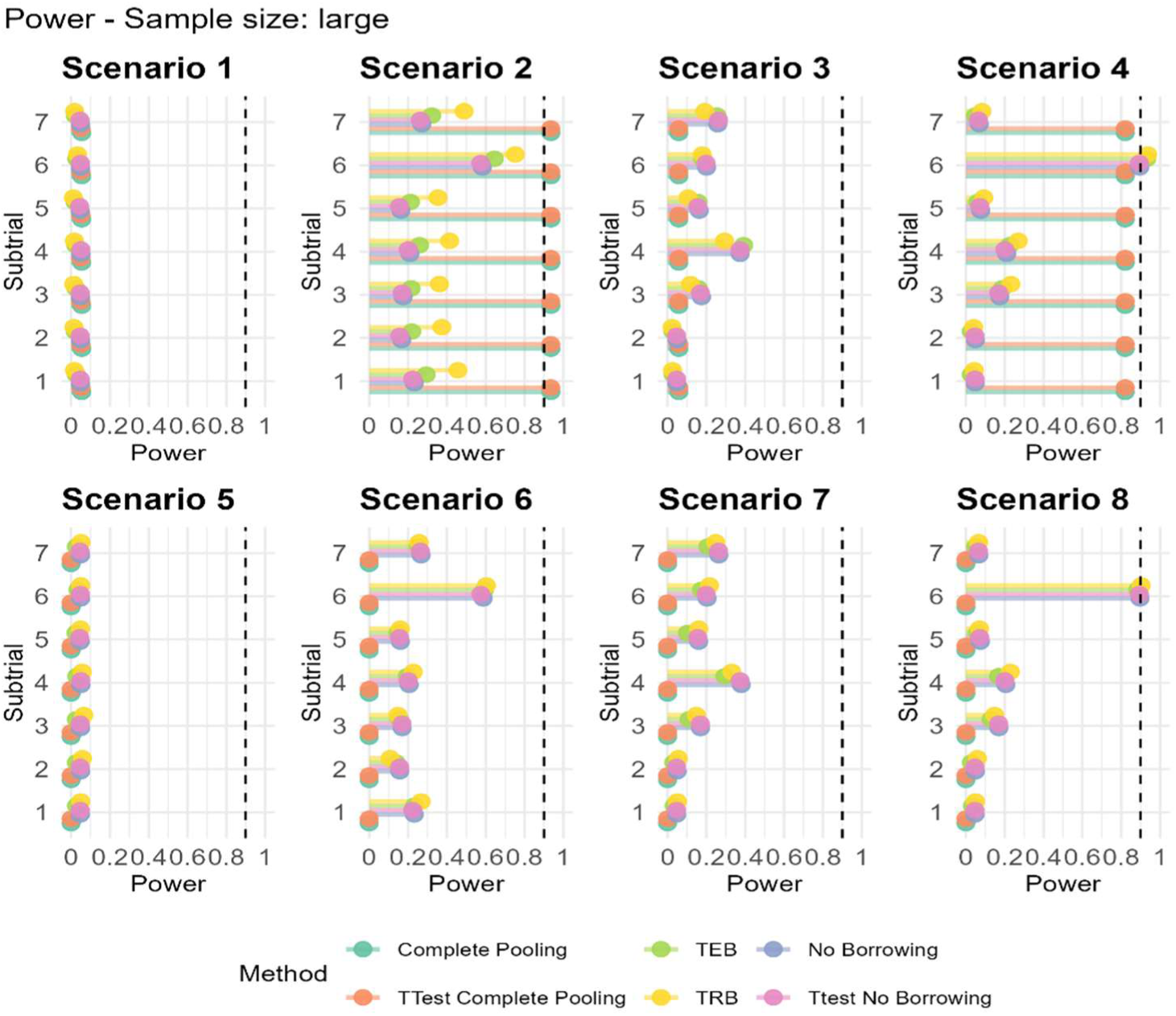
**A Power and Type 1 error for large sample size**

**Figure 10.**
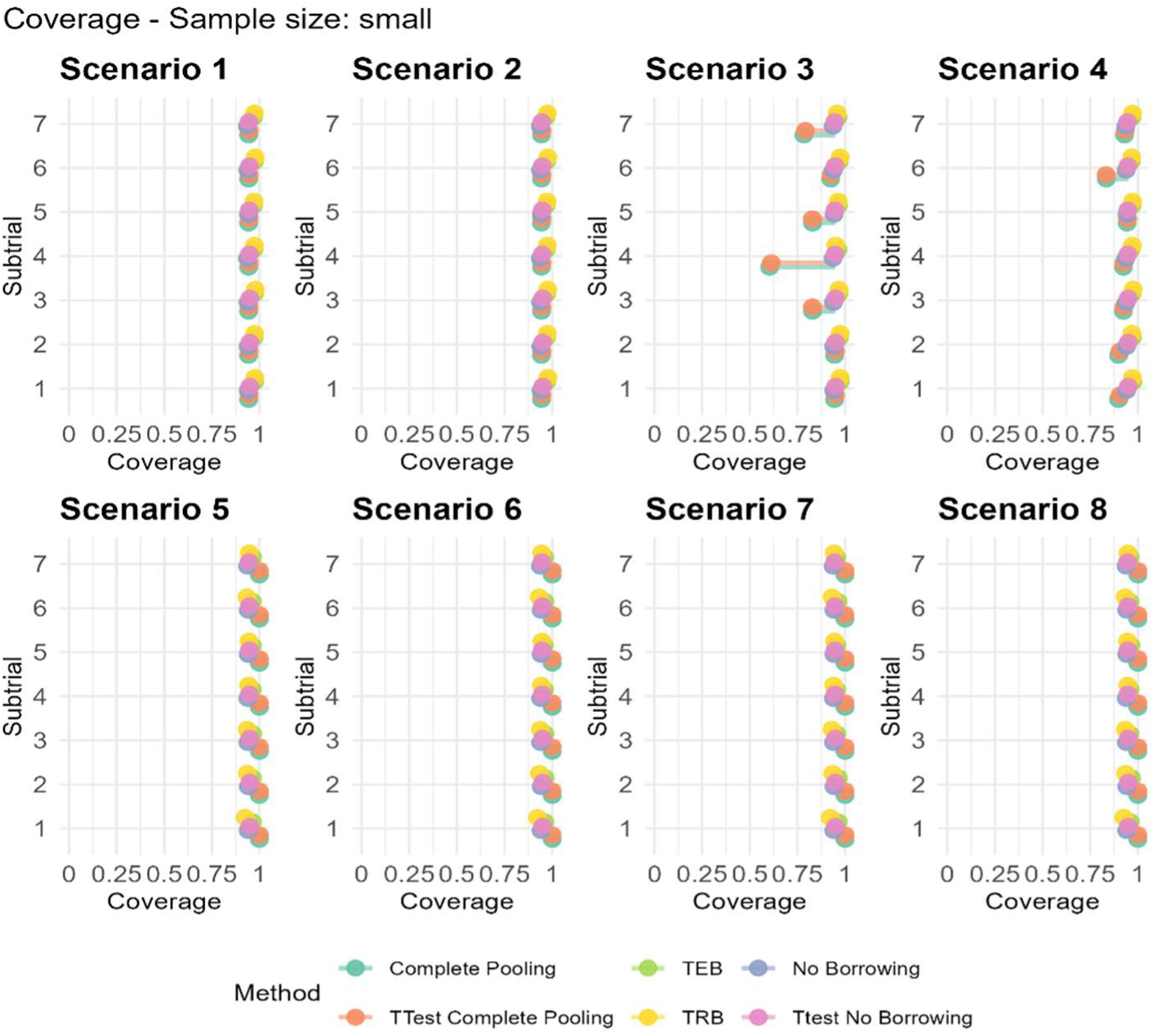
**A Coverage for small sample size**

**Figure 11.**
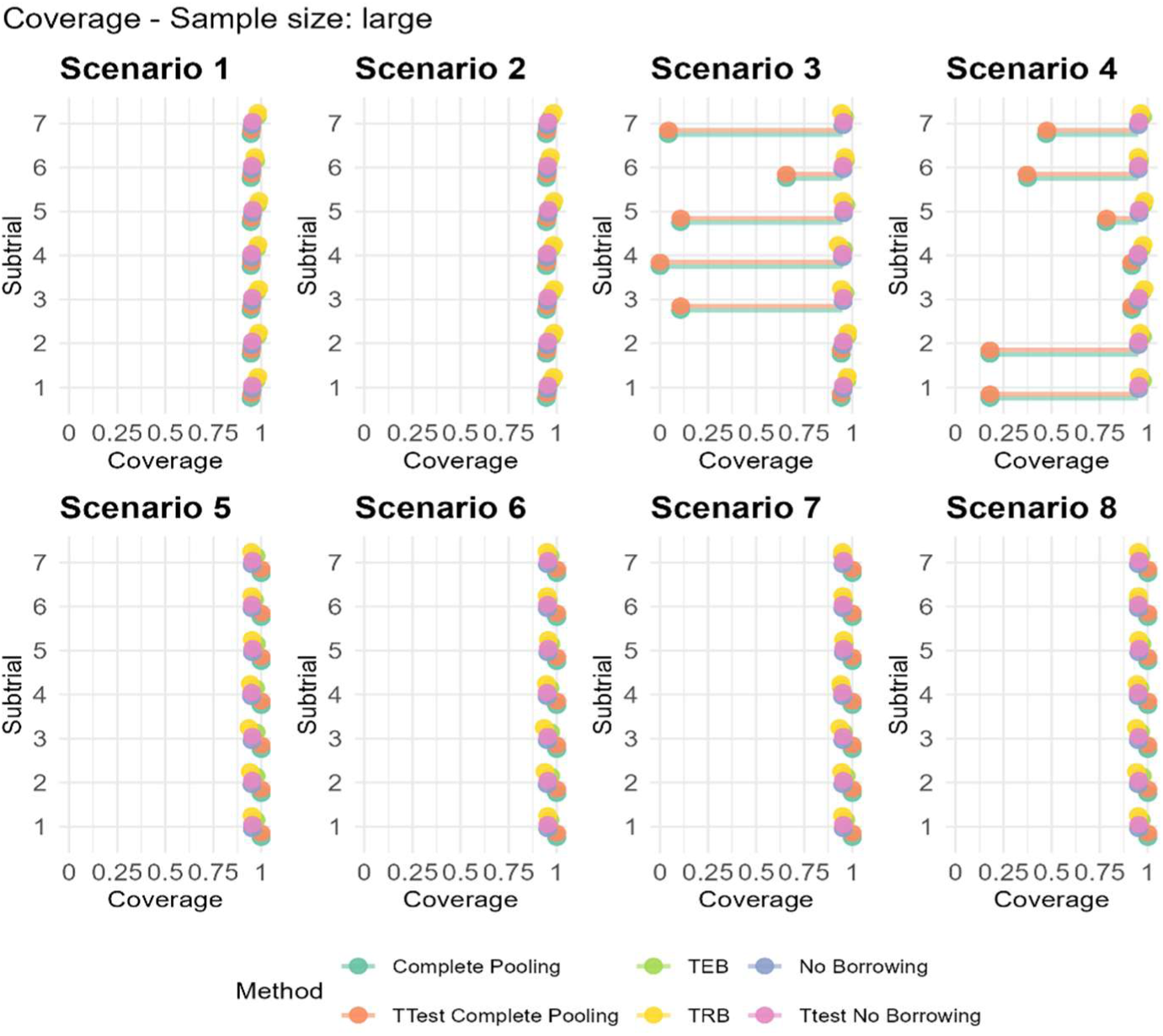
A Coverage for large sample size

**Figure 12.**
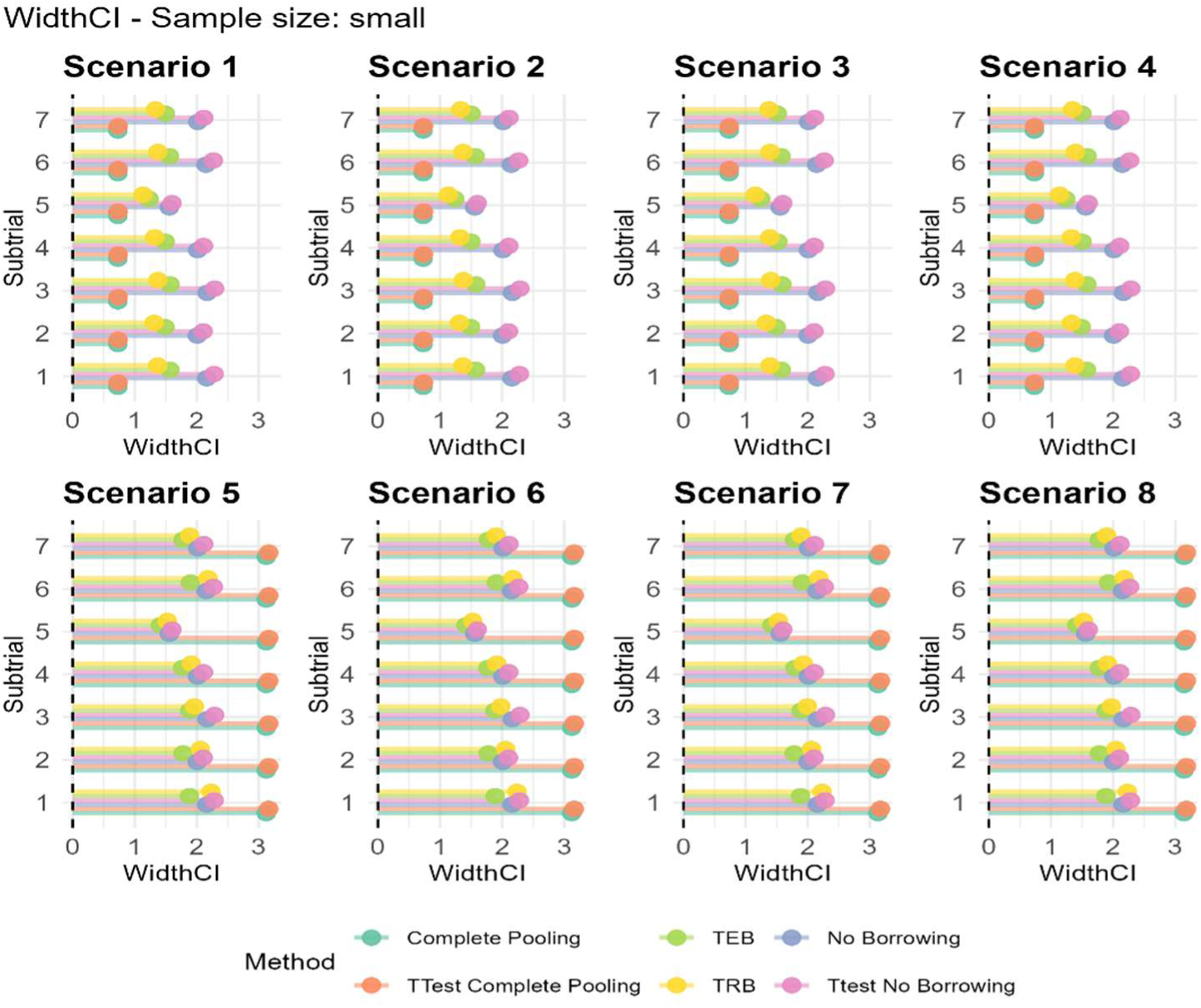
**A Width of CI for small sample size**

**Figure 13.**
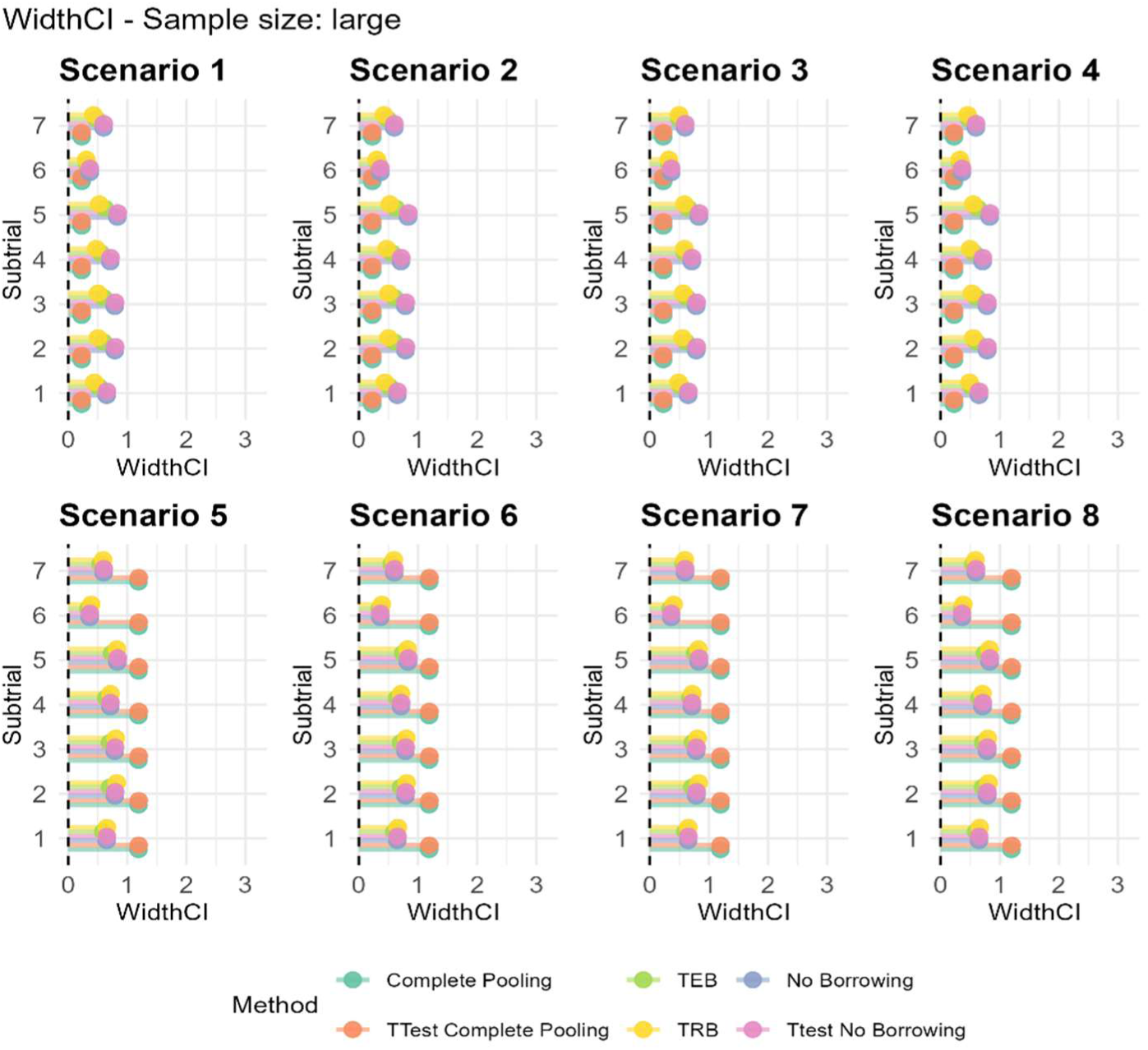
**A Width of CI for large sample size**

**Figure 14.**
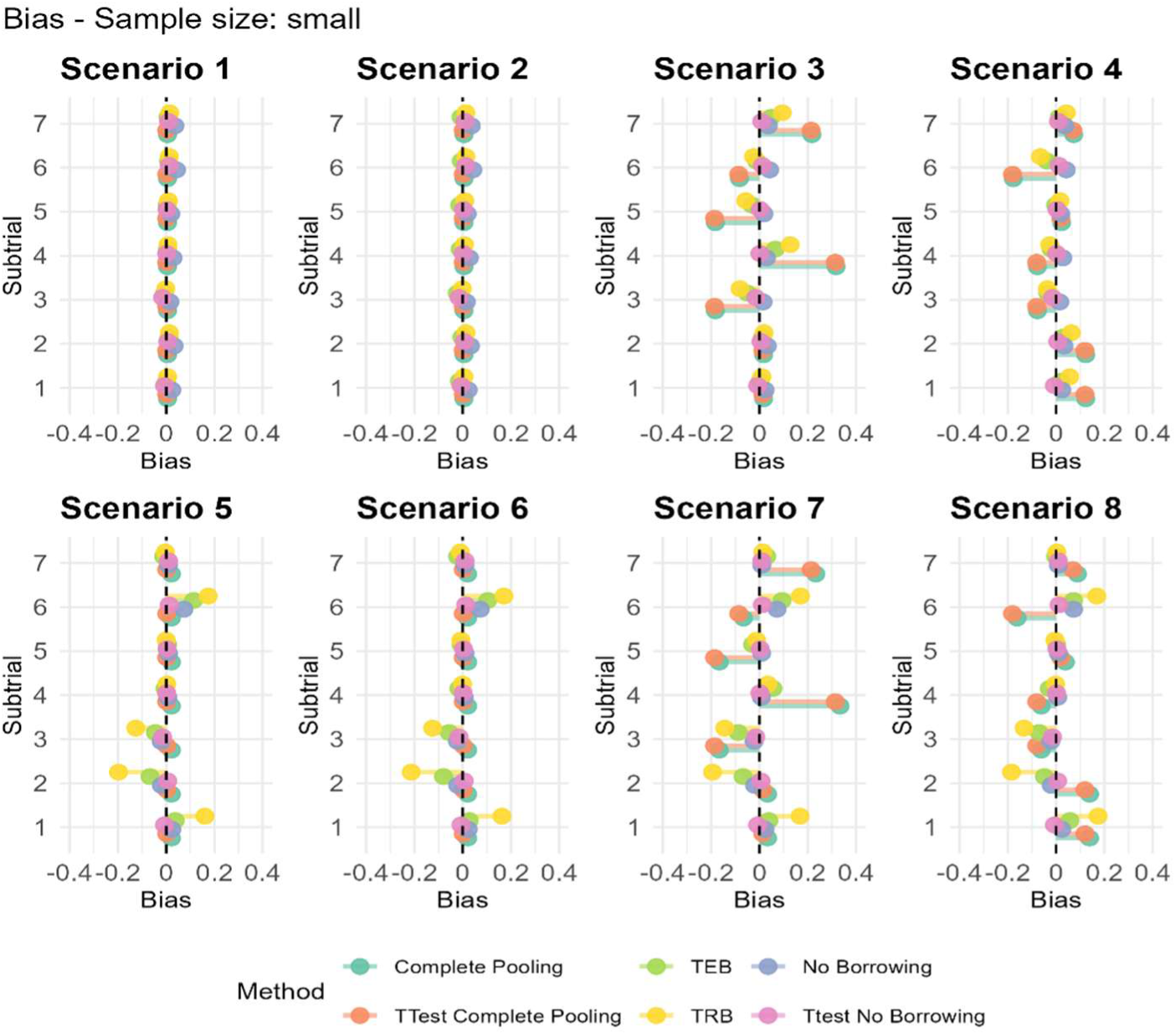
**A Bias for small sample size**

**Figure 15.**
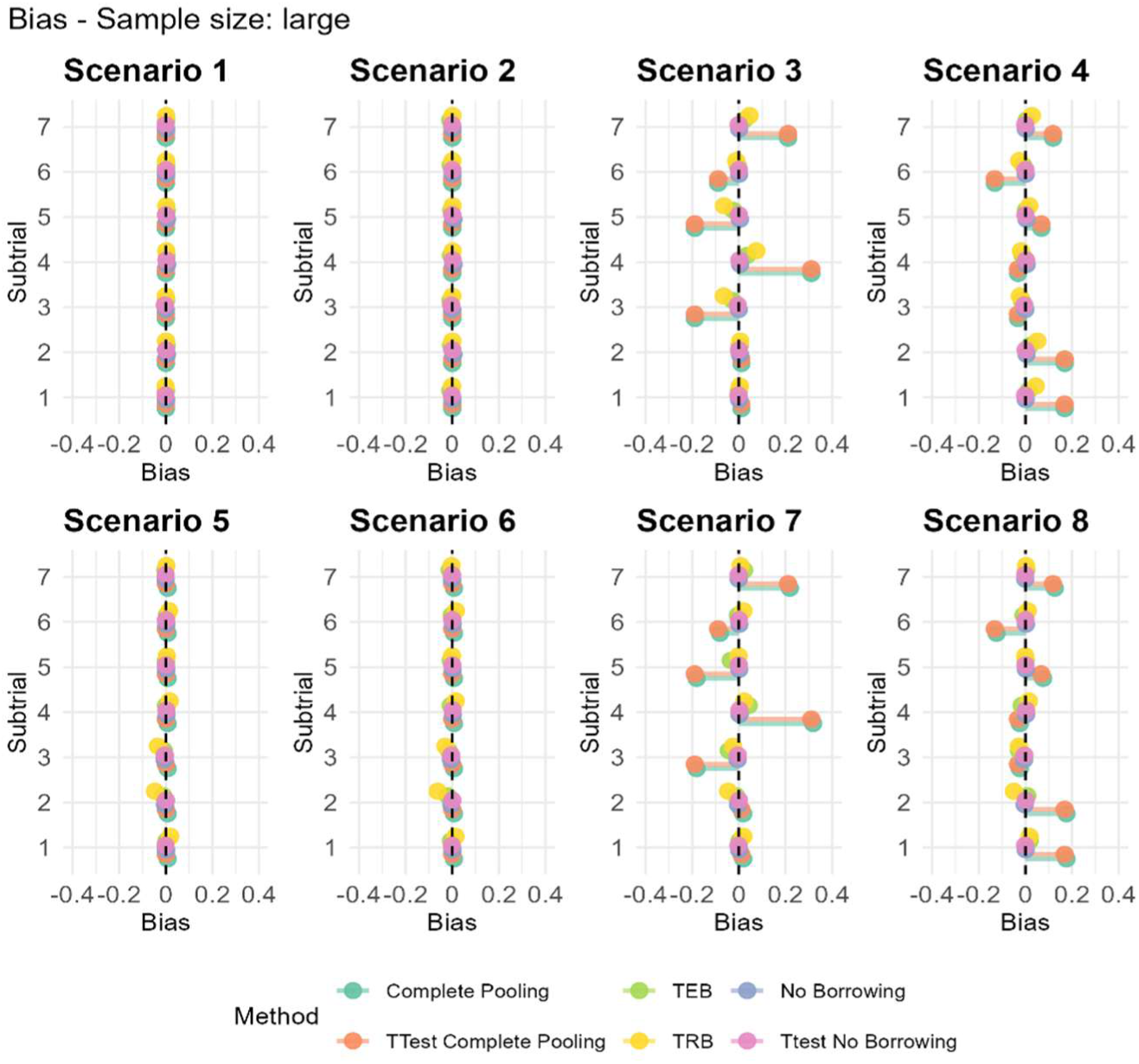
**A Bias for large sample size**

**Figure 16.**
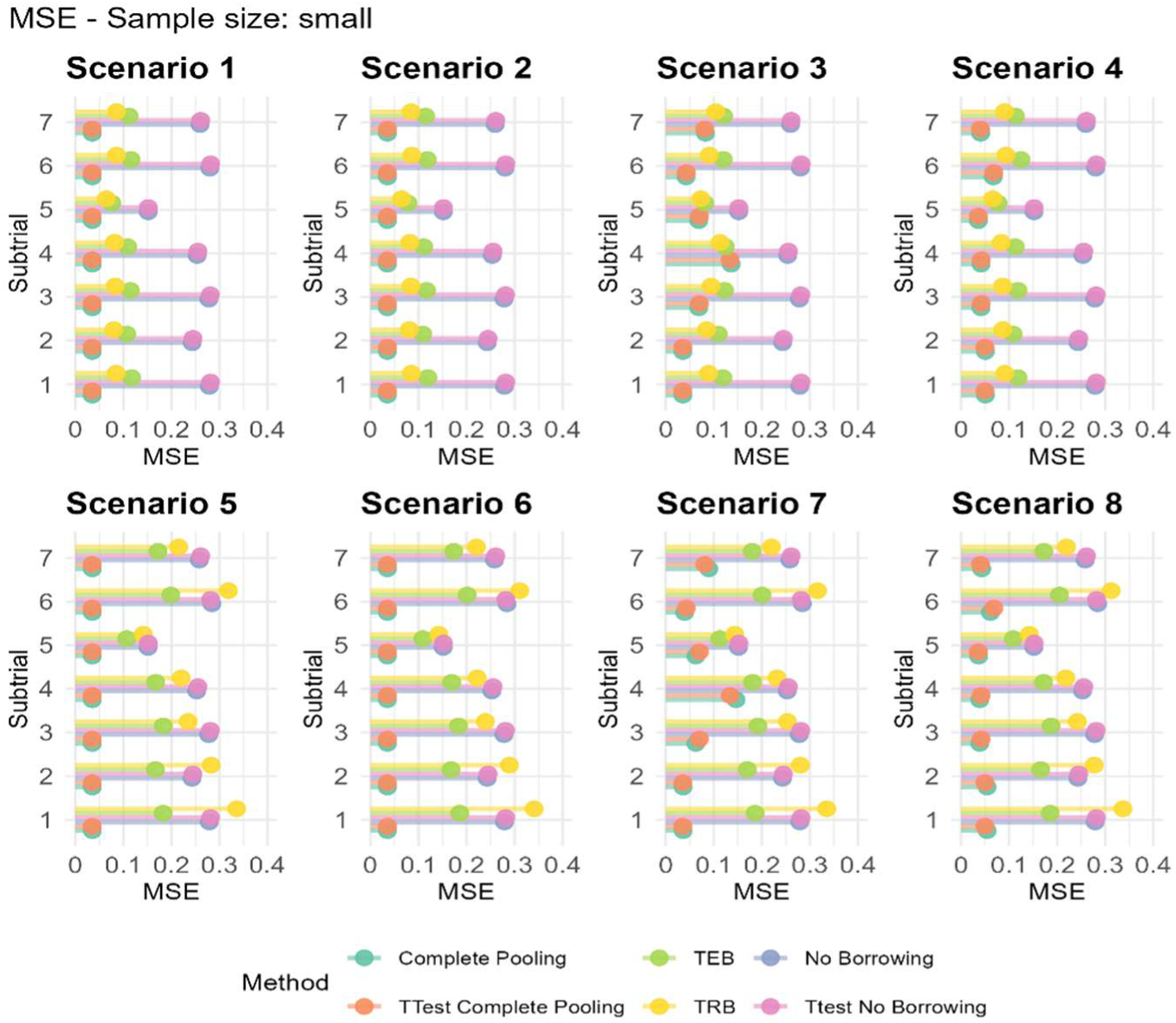
**A MSE for small sample size**

**Figure 17.**
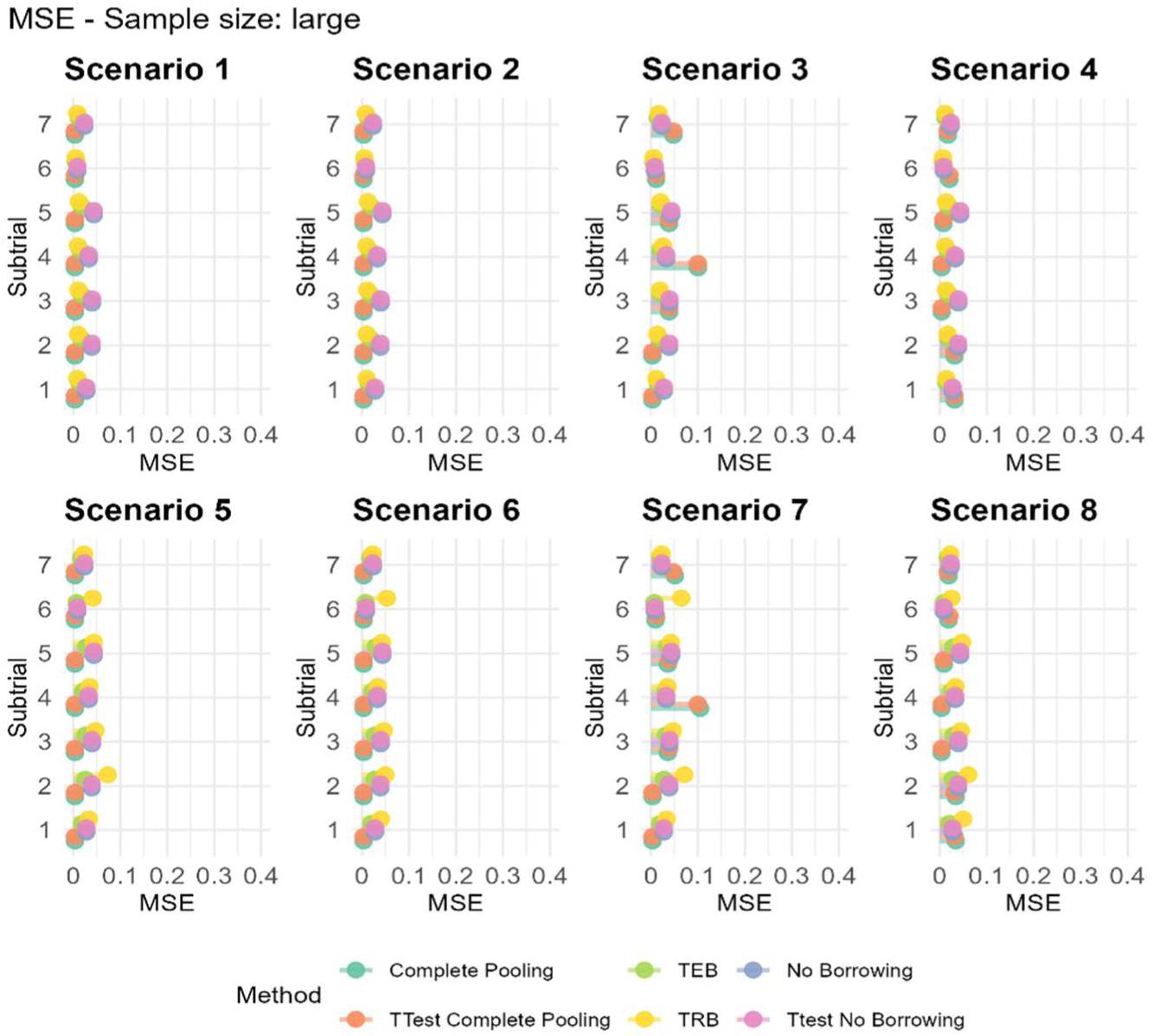
**A MSE for large sample size**

**Figure 18.**
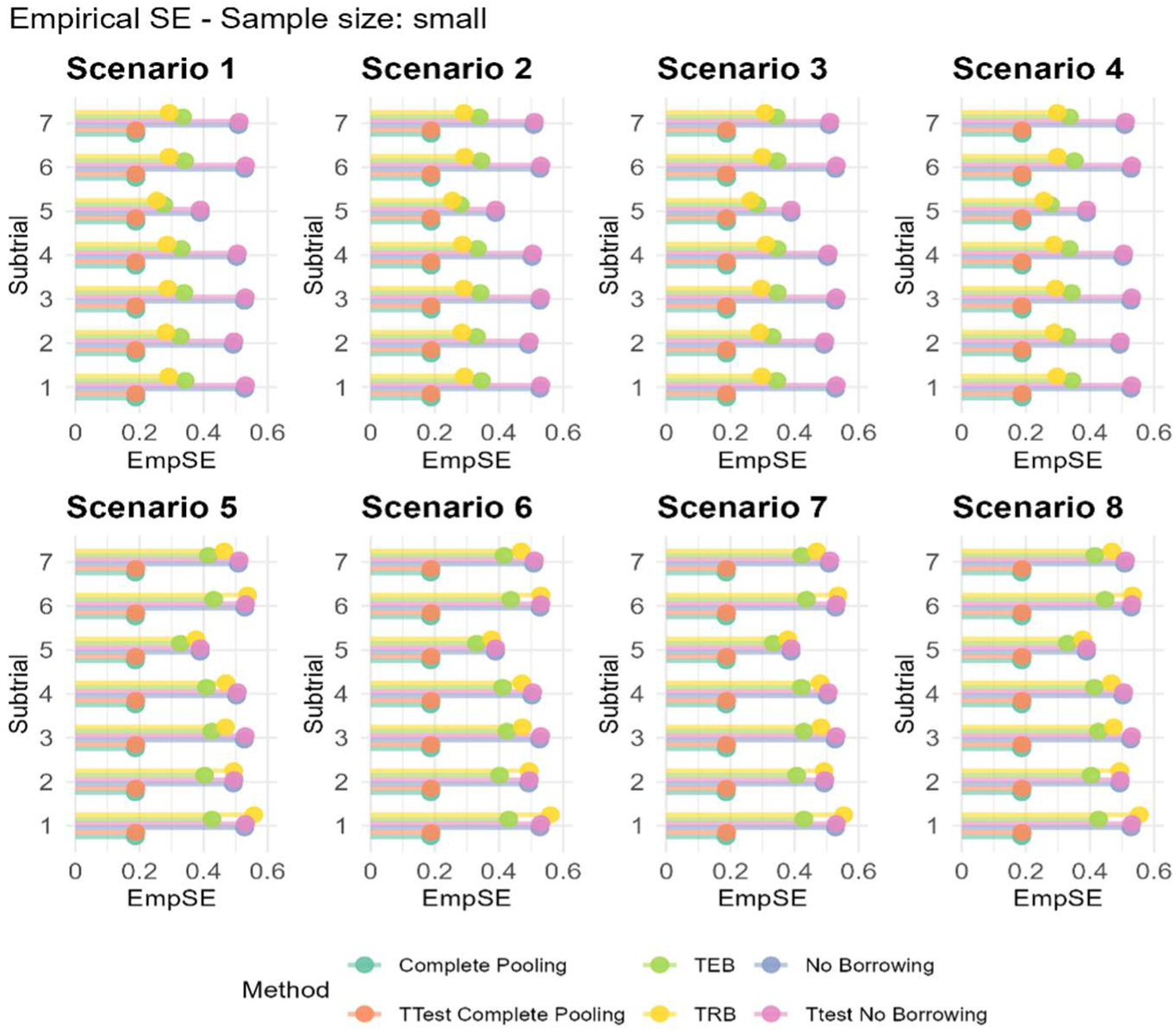
**A Empirical SE for small sample size**

**Figure 19.**
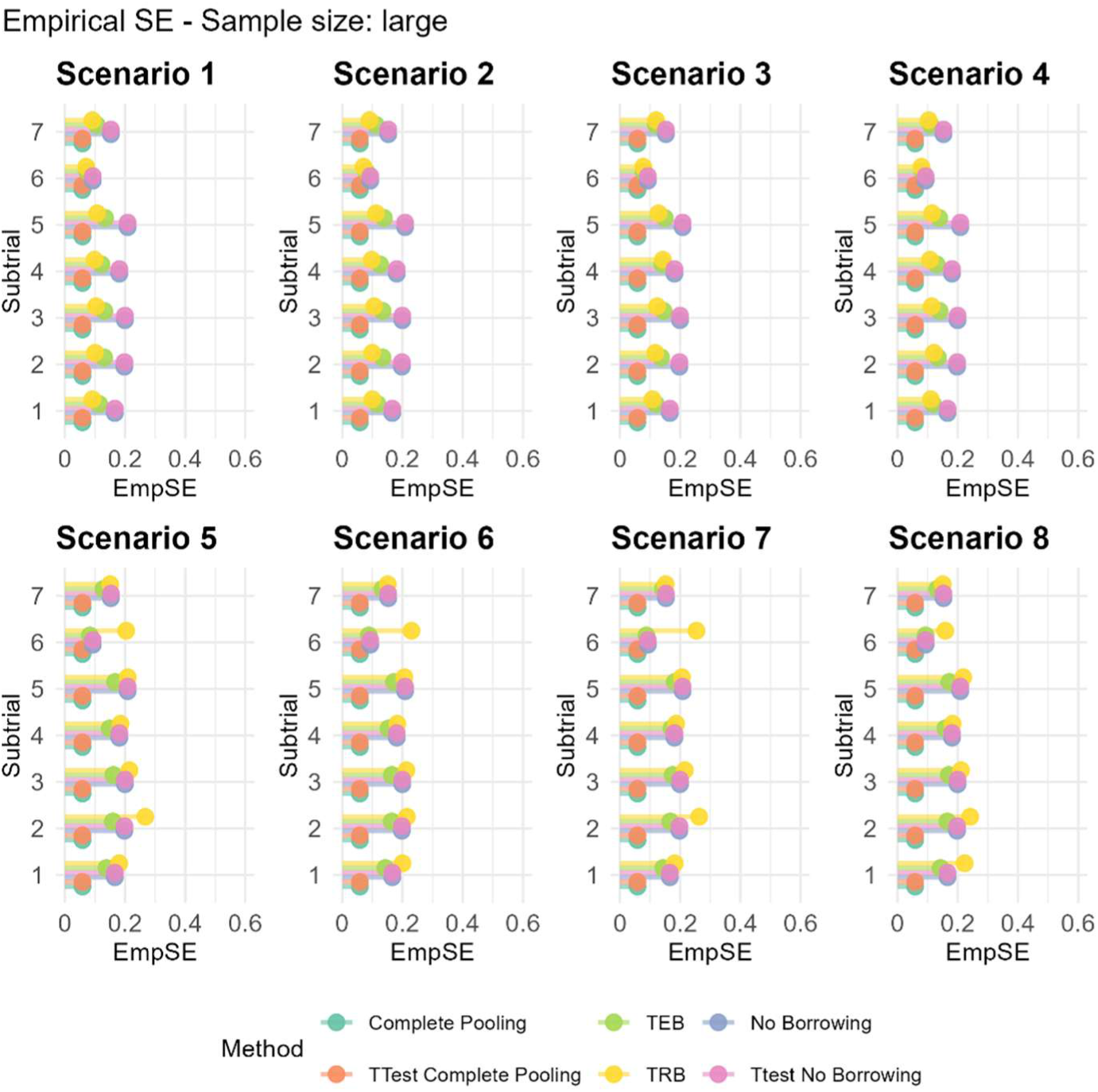
**A Empirical SE in large sample size**

## Discussion

Our simulation study was motivated by the need to reduce research waste by allowing patients with heterogeneous symptoms to be evaluated within a common trial framework. Our earlier systematic review demonstrated inefficiencies in current trial designs in gynaecology, while our network meta-analysis highlighted challenges for evidence synthesis arising from variation in outcome measurement and the scales used across trials. Together, these findings motivate the development of a trial design in gynaecology that make more efficient use of the available patient population, whilst ensuring that outcomes important to patients, such as their most bothersome symptom, are appropriately utilised.

We evaluated the performance of basket trials with differing levels of information borrowing, in a gynaecology setting where patients with the same underlying condition are allocated to subtrials based upon their “most bothersome symptom” and randomised to receive either an intervention or control. We compared several Bayesian borrowing strategies with two extreme approaches: no borrowing and complete pooling. These comparators were chosen to reflect common trial designs in gynaecology identified in a previous systematic review (5). The no borrowing approach represents the traditional symptom-specific randomised controlled trial (RCT), in which eligibility is restricted to patients presenting with a particular symptom. Although clinically targeted, these trials often recruit relatively small numbers of participants and consequently have limited statistical power. In contrast, complete pooling reflects studies that recruit broader patient populations regardless of symptom presentation and analyse outcomes across all participants regardless of relevance to the patient. While this approach maximises the pool of eligible participants, it may introduce inefficiencies and bias by combining information across clinically heterogeneous groups.

Recent recommendations by Broglio et al. suggest that borrowing between subtrials should only be considered when three conditions are satisfied: (1) the disease populations are sufficiently related; (2) there is a sound biological rationale for expecting similar treatment effects in each subtrial, and (3) the measurement and relevance of the outcome assessment is the same in each subtrial These recommendations informed the design of our hypothetical basket trial. All participants in our simulations shared the same underlying gynaecological condition, with subtrials defined according to participants’ most bothersome symptom, thereby providing a setting in which the disease populations were related, satisfying recommendation (1). Although assessment of the intervention’s mechanism of action ultimately requires clinical expertise, a common intervention targeting multiple symptoms arising from the same condition provides a plausible rationale for similar treatment effects across subtrials. We nevertheless varied treatment effects across simulation scenarios to assess the impact of departures from this assumption, therefore testing recommendation (2). Finally, the same outcome assessment was applied consistently across all subtrials, satisfying recommendation (3). Together, these scenarios allowed us to investigate how the suitability and performance of Bayesian borrowing changes across clinically plausible subtrial settings, including differences in treatment effects, control-group means and sample sizes.

In our simulation, we observed that Bayesian and frequentist implementations of the No Borrowing approach yielded similar but not identical results. This discrepancy reflects the inherent methodological difference between the two frameworks: the Bayesian approach incorporates prior information (even when weakly informative), while the frequentist approach relies solely on the likelihood of the observed data. In contrast, Bayesian and frequentist Complete Pooling approaches produced little to no difference in results across all performance measures.

While borrowing approaches improved power relative to No Borrowing in scenarios with homogeneous treatment effects, gains were reduced or inconsistent under heterogeneity, highlighting the impact of shrinkage-induced bias. For Complete Pooling power was reduced in scenarios with differing control group means, as the increased between-subtrial variability was not accounted for in the analysis (i.e. the adjustment for subtrial), as in other borrowing methods. Coverage was broadly similar across all methods and scenarios, with values of at least 95%. However, Complete Pooling (CP) produced notably poor coverage in scenarios with heterogeneous treatment effects (Scenarios 3 and 4). These results demonstrate that taking the overall pooled estimate, using Complete Pooling, will not provide a good estimate off the subtrial-specific effects when they are very heterogeneous. From a practical perspective, the results of our simulations study suggest that while adequate sample size remains important for achieving sufficient power and precision, the choice between methods is unlikely to depend strongly on sample size, as their comparative behaviour was largely consistent across all settings examined.

Overall, the performance metrics for our simulation paint a nuanced picture. Taken together, the findings suggest that no single borrowing strategy is universally optimal. In fact, the performance of each borrowing strategy depends heavily on the degree and nature of between-subtrial heterogeneity. Complete Pooling offers potential efficiency gains when subtrials are sufficiently homogeneous; otherwise, it introduces meaningful bias. Moreover, when baseline responses differ substantially between subtrials, the assumption made by CP that there is a common baseline response can result in highly conservative inference. Conversely, analysing subtrials independently is potentially preferable when heterogeneity in treatment effect is high. However, this method lacks statistical power when treatment effects are similar. Adaptive borrowing approaches (TEB, TRB) provide a compromise by allowing the extent of information sharing to depend on the observed similarity between subtrials. Among these methods, TEB appeared to be more robust than TRB when control arm responses differ across subtrials, suggesting that it may be the preferable approach in settings where baseline heterogeneity cannot be ruled out.

These findings support the recommendations of Broglio et al, that no single borrowing strategy should not be applied routinely but instead selected according to the plausibility of exchangeability between subtrials (16). Clinical understanding of disease mechanisms, anticipated treatment responses and patient characteristics should therefore inform both the decision to use a basket design and the choice of borrowing strategy. Our simulations reinforce this principle: borrowing improved efficiency when treatment effects were similar but introduced bias when similarity assumptions were violated. Consequently, careful assessment of exchangeability during trial design is essential, and adaptive borrowing methods are likely to be preferable whenever uncertainty exists regarding between-subtrial similarity.

An important implication of this work concerns the role of basket trials within gynaecological research. One alternative to a basket design would be to recruit all patients into a single trial and analyse a universally applicable outcome measure. However, this risks overlooking symptom-specific treatment effects that may be clinically important. The basket trial framework offers a compromise by allowing patients with the same underlying condition but different predominant symptoms to be studied within a single protocol while preserving symptom-specific analyses. When borrowing assumptions are appropriate, this approach can improve efficiency without ignoring clinically relevant heterogeneity. Future methodological work should investigate settings in which different outcomes are measured across subtrials. Additionally, further work exploring different outcome types would be beneficial as complete pooling is unlikely to be appropriate in these circumstances and alternative borrowing strategies may be required.

Basket trials represent an important advancement towards precision medicine, and an opportunity to improve the efficiency of research. However, they also introduce several methodological and practical challenges. Appropriate prior specification, control of false positive and false negative findings, and decisions regarding the extent of information sharing require careful statistical consideration, while coordinating multiple subtrials may increase logistical complexity and cost. Nevertheless, when supported by a strong clinical rationale, basket trials have the potential to evaluate treatments across heterogeneous symptom presentations more efficiently than conducting multiple independent trials.

The publication of the UK government’s “Women’s Health Strategy for England” has highlighted the need for more high-quality research into women’s health, aiming to improve health outcomes and data usage (17). With this in mind, developing robust statistical methodology for efficient, patient-centred gynaecological trials is particularly timely. Our findings suggest that basket trial designs, combined with carefully selected Bayesian borrowing strategies, represent a promising approach for evaluating interventions across patients with shared underlying conditions but differing symptom profiles, provided that assumptions regarding exchangeability are carefully considered during trial design.

## Conclusions

Complete Pooling, analogous to a single large clinical trial pooling all patients, performs well where the subtrials are highly homogenous but becomes problematic when meaningful between-subtrial heterogeneity is present. In contrast, No Borrowing (representing the conventional approach of conducting multiple small, independent trials), avoids the bias introduced by inappropriate pooling but sacrifices efficiency, yielding estimates with more uncertainty. Neither extreme is therefore universally satisfactory.

Adaptive borrowing approaches offer a compromise: treatment effect borrowing (TEB) is generally more robust across a range of scenarios, while treatment response borrowing (TRB) performs competitively only when homogeneity in the control arm can be reasonably assumed. Therefore, we conclude with caution that borrowing information in a randomised basket trial setting offers efficiency gains that are not currently seen in gynaecological trials. Basket trials should only be considered if there is clinical and biological justification.

## Appendices

## Data Availability

All data produced in the present work are contained in the manuscript. All code used in this study for both R and JAGS is available at https://github.com/katiestones/basket-sim

